# The tryptophan catabolite or kynurenine pathway in major depressive and bipolar disorder: a systematic review and meta-analysis

**DOI:** 10.1101/2022.06.13.22276359

**Authors:** Abbas F. Almulla, Yanin Thipakorn, Asara Vasupanrajit, Ali Abbas Abo Algon, Chavit Tunvirachaisakul, Ashwan Abdulzahra Hashim Aljanabi, Gregory Oxenkrug, Hussein K. Al-Hakeim, Michael Maes

## Abstract

**Background:** There is now evidence that affective disorders including major depressive disorder (MDD) and bipolar disorder (BD) are mediated by immune-inflammatory and nitro-oxidative pathways. Activation of these pathways may be associated with activation of the tryptophan catabolite (TRYCAT) pathway leading to depletion of tryptophan (TRP) and increases in tryptophan catabolites (TRYCATs).

**Aims:** To systematically review and meta-analyze TRP, its competing amino-acids (CAAs) and TRYCAT data in MDD and BD.

**Methods:** This review searched PubMed, Google Scholar and SciFinder and included 121 full-text articles and 15470 individuals, including 8024 MDD/BD patients and 7446 healthy controls.

**Results:** TRP levels (either free and total) and the TRP/CAAs ratio were significantly decreased (p<0.0001) in MDD/BD as compared with controls with a moderate effect size (standardized mean difference for TRP: SMD=-0.513, 95% confidence interval, CI: -0.611; -0.414; and TRP/CAAs: SMD=-0.558, CI: -0.758; -0.358). Kynurenine (KYN) levels were significantly decreased in patients as compared with controls with a small effect size (p<0.0001, SMD= -0.213, 95%CI: -0.295; -0.131). These differences were significant in plasma (p<0.0001, SMD=-0.304, 95%CI: -0.415, -0.194) but not in serum (p=0.054) or the central nervous system (CNS, p=0.771). The KYN/TRP ratio, frequently used as an index of indoleamine-dioxygenase (IDO) activity, and neurotoxicity indices based on downstream TRYCATs were unaltered or even lowered in MDD/BD.

**Conclusions:** Our findings revealed that MDD/BD are accompanied by TRP depletion without IDO and TRYCAT pathway activation. Lowered TRP availability is probably the consequence of lowered serum albumin during the inflammatory response in affective disorders.

## Introduction

There is now robust evidence that activation of the immune-inflammatory response system (IRS) and the compensatory immune-regulatory system (CIRS) play an essential role in the pathophysiology of major depressive (MDD) and bipolar (BD) disorder (Maes and Carvalho 2018, Almulla and Maes 2022). Both disorders are characterized by elevated production of macrophage M1 and T helper (Th)1 cytokines including interleukin (IL)-1β, IL-6, IL-8, interferon (IFN)-γ, tumor necrosis factor (TNF)-α, whereas CIRS activation is indicated by elevated levels of anti-inflammatory products including Th-2 and Tregulatory (Treg) cytokines including IL-4 and IL-10 (Maes and Carvalho 2018). Both disorders are also accompanied by an acute-phase (AP) response with increased levels of positive AP proteins (APPs) such as haptoglobin and lowered levels of negative APPs such as albumin (Maes 1993). Moreover, the activated IRS pathways in affective disorders are associated with activation of nitro-oxidative pathways with increased reactive oxygen and nitrogen species (RONS), and consequent lipid peroxidation and protein oxidation (Maes, Galecki et al. 2011, Maes 2022). Activation of IRS and nitro-oxidative pathways are associated with the key features of affective disorders, including severity of illness, staging (reoccurrence of episodes) and suicidal behaviors including suicidal ideation and attempts (Vasupanrajit, Jirakran et al. 2021, Maes 2022, Maes, Rachayon et al. 2022). The current theory is that the neurotoxic effects of M1 and Th-1 cytokines and RONS cause neuro-affective toxicity with dysfunctions in brain connectome pathways that lead to staging and the phenome of MDD/BD (Maes 2022, Maes, Rachayon et al. 2022).

Activation of IRS and nitro-oxidative pathways has a number of major detrimental consequences including depletion of tryptophan (TRP) in peripheral blood and increases in levels of neurotoxic tryptophan catabolites (TRYCATs) (Maes, Leonard et al. 2011, Maes 2015). Since tryptophan binds tightly to albumin, the decreased levels of albumin during the acute phase or IRS response in affective disorders may result in a decrease in total TRP levels in peripheral blood (Maes, Leonard et al. 2011). Moreover, products of the IRS and nitro-oxidative stress response during MDD/BD may activate indoleamine-2,3-dioxygenase (IDO), the rate limiting enzyme of the TRP catabolite (TRYCAT) pathway, which may cause increased TRYCATs production and lower TRP thereby diverting TRP from serotonin synthesis (Maes, Leonard et al. 2011) (**Figure 1)**. These activators include reactive oxygen species (ROS), IL-1β, TNF-α, IFN-α and IFN-γ (Maes, Leonard et al. 2011) and lipopolysaccharides (LPS) generated by translocation of Gram-negative bacteria (Maes, Kubera et al. 2008). While IDO is active in immune cells (macrophages and dendritic cells) and brain cells (e.g. astrocytes), tryptophan-2,3-dioxygenase (TDO) is activated by glucocorticoids and is expressed primary in the liver where it converts TRP to the same TRYCATs (Maes, Minner et al. 1991, Maes, Leonard et al. 2011).

**Figure 1:**
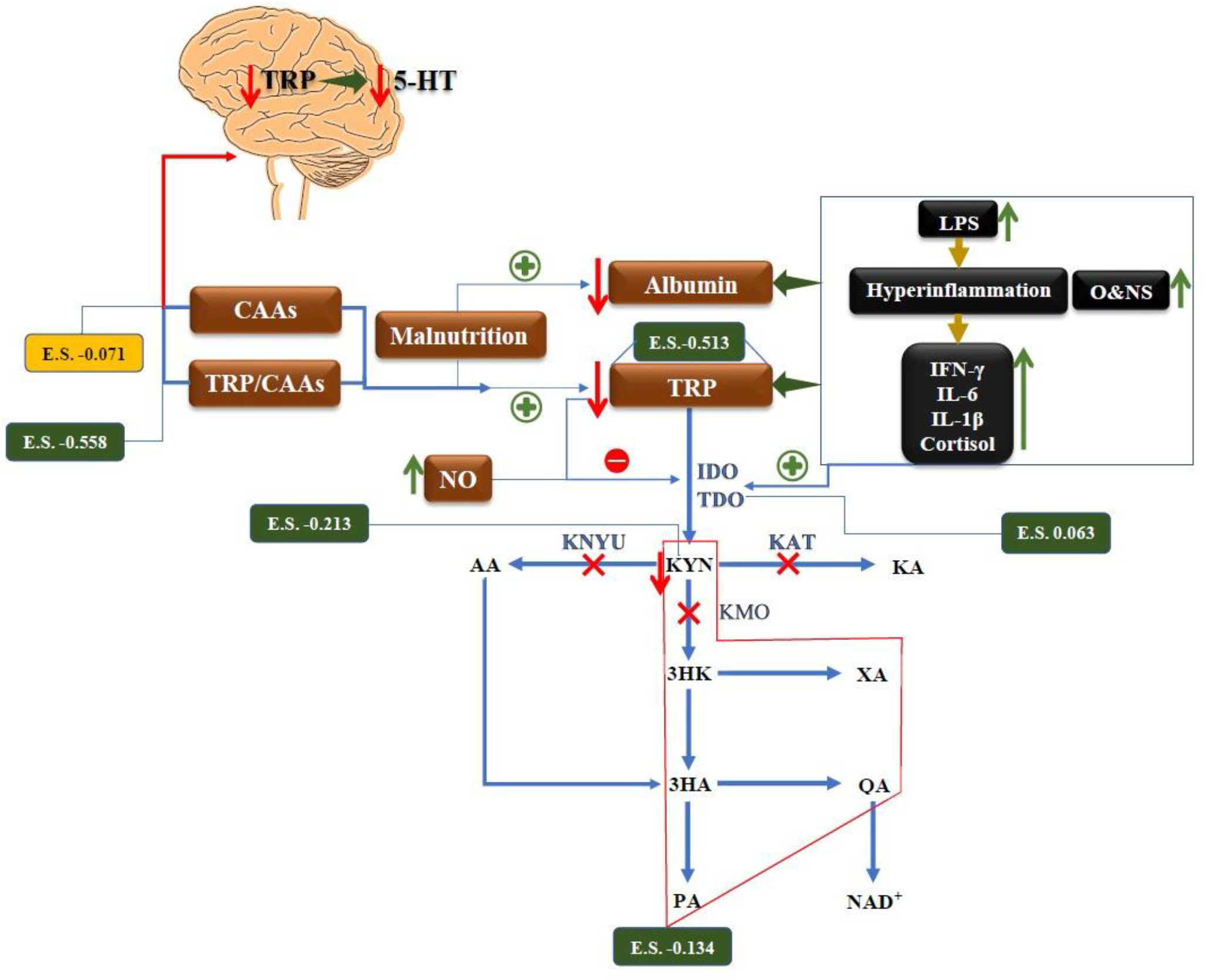
A Summary of tryptophan catabolite (TRYCAT) pathway in affective disorders. E.S.: Effect size, TRYCAT: Tryptophan catabolite, IFN-γ: Interferon-Gamma, IL-6: Interleukin 6, IL-1β: Interleukin-1 beta, O&NS: Oxidative and nitrosative stress, NO: Nitric Oxide, 5-HT: 5-Hydroxytryptamine, LPS: Lipopolysaccharides, CNS: Central nervous system, IDO: Indoleamine 2,3 dioxygenase, TDO: Tryptophan 2,3-dioxygenase, KAT: Kynurenine Aminotransferase, KMO: Kynurenine 3-monooxygenase, KYNU: Kynureninase, TRP: Tryptophan, KYN: Kynurenine, KA: Kynurenic Acid, 3HK: 3-Hydroxykynurenine, AA: Anthranilic Acid, XA: Xanthurenic Acid, 3HA: 3-Hydroxyanthranilic Acid, PA: Picolinic Acid, QA: Quinolinic Acid.

Overall, stimulation of the TRYCAT pathway has CIRS functions including anti-inflammatory and antioxidant effects in part through reductions in TRP, which results in negative immune-regulatory, antiproliferative and antimicrobial effects, and elevations in TRYCATs which have anti-inflammatory and antioxidant effects (Maes, Leonard et al. 2011, Almulla and Maes 2022). For example, kynurenic acid (KA), xanthurenic acid (XA) and quinolinic acid (QA) may exert anti-inflammatory effects through their ability to decrease the IFN-γ/IL-10 ratio (Maes, Mihaylova et al. 2007, Maes 2015, Almulla and Maes 2022). On the other hand, increases in downstream TRYCATs may cause neurotoxicity: kynurenine (KYN), QA, picolinic acid (PA) and XA have neurotoxic effects, and 3-hydroxyanthranilinc acid (3HA), 3-hydroxykynurenine (3HK) and QA may induce oxidative stress (Santamaría, Galván-Arzate et al. 2001, Smith, Smith et al. 2009, Reyes Ocampo, Lugo Huitron et al. 2014).

Previous research (Maes, Jacobs et al. 1990, Maes, De Backer et al. 1995, Marx, McGuinness et al. 2021) and meta-analysis (Ogawa, Fujii et al. 2014) revealed that patients with MDD and BD show reduced TRP levels. In mood disorders, low TRP has been reported to be a biomarker of IRS activation and the acute phase response (Maes 2015), whilst some but not all reports show increased TRYCATs levels in MDD/BD. Ogyu et al. showed that drug-free depressed patients have diminished levels of KYN and KA and increased QA levels (Ogyu, Kubo et al. 2018). Furthermore, Arnone et al. reported that MDD patients demonstrated lowered KYN level compared with healthy controls (Arnone, Saraykar et al. 2018). Marx et al. found that MDD was accompanied by decreased TRP, KYN and KA levels and that BD patients showed reduced TRP and KA levels (Marx, McGuinness et al. 2021). Hebbrecht et al. reported lowered peripheral TRP, KYN, KA levels in BD patients compared to healthy controls (Hebbrecht, Skorobogatov et al. 2021).

TRYCATs with neurotoxic activities including KYN and 3-HK play a key role in IFN-α induced major depression (Bonaccorso, Marino et al. 2001, Maes, Bonaccorso et al. 2001). Indeed, the onset of these IFN-α-induced depressive symptoms is more strongly associated with the production of KYN (a neurotoxic TRYCAT) and an increased ratio of KYN to KA, a neuroprotective TRYCAT, than with lowered TRP levels (Bonaccorso, Marino et al. 2001, Maes, Bonaccorso et al. 2001, Bonaccorso, Marino et al. 2002, Wichers, Koek et al. 2005, Maes 2015). By inference, it was thought that also in MDD/BD, which is characterized by lowered plasma/serum tryptophan, increased neurotoxicity due to IDO stimulation could be the major culprit (Bonaccorso, Marino et al. 2002).

Nevertheless, the abovementioned meta-analyses did not include the levels of free or total TRP and their competing amino acids (CAAs), namely valine, tyrosine, leucine, isoleucine and phenylalanine, in order to evaluate the TRP/CAAs ratio which is a more adequate index reflecting TRP availability to the brain than plasma/serum TRP (Lucini, Lucca et al. 1996). In addition, these meta-analyses did not provide sufficient evidence to determine whether affective disorders are related with alterations in downstream neurotoxic TRYCATs due to activated IDO activity. Finally, we recently discovered that the serum and plasma TRYCAT results in schizophrenia are dissociated from central nervous system (CNS) findings, and that there are significant discrepancies in the correlations between schizophrenia and KYN in serum versus plasma (Almulla, Vasupanrajit et al. 2022).

Hence, we conducted a systematic review and meta-analysis to comprehensively examine free/total TRP, the sum of CAAs and the TRP/CAAs ratio along with indices of IDO, Kynurenine aminotransferase (KAT) and Kynurenine 3-monooxygenase (KMO) enzyme activities, and the levels of downstream neurotoxic TRYCATs in both MDD and BD in CNS, serum and plasma.

## Materials and methods

We conducted the current meta-analysis to examine the peripheral (serum and plasma) and central (cerebrospinal fluid, CSF and brain) levels of TRP, KYN, KA and 3HK as well as some ratios namely KYN/TRP which reflects the IDO enzyme activity, KA/KYN (KAT enzyme activity and 3HK/KYN (KMO enzyme activity) along with the neurotoxic TRYCAT composite (KYN+3HK+3HA+QA+XA+PA) in patients with MDD and BD. In the same patients we also assessed peripheral level of CAAs and TRP/CAAs ratio.

The methodology of this study was based on the guidelines of a) Preferred Reporting Items for Systematic Reviews and Meta-Analyses (PRISMA) 2020 (Page, McKenzie et al. 2021), b) Cochrane Handbook for Systematic Reviews and Interventions (Higgins JPT 2019), c) the Meta-Analyses of Observational Studies in Epidemiology (MOOSE).

### Search strategy

The 10th of January 2022 marked the beginning of our examination of the electronic databases PubMed/MEDLINE, Google Scholar, and SciFinder. To collect all the publications pertaining to TRP and TRYCATs in affective disorders, we used the keywords and mesh terms provided in Table 1 of the Electronic supplementary file (ESF). The database search concluded at the end of March 2022. Nevertheless, we verified the reference lists of all eligible papers and previous meta-analyses to prevent the omission of pertinent publications.

**Table 1.** The outcomes and number of patients with affective disorders and healthy control along with the side of standardized mean difference (SMD) and the 95% confidence intervals with respect to zero SMD.

| Outcome profiles | n<br>studies | Side of 95% confidence intervals |  |  |  | Patient<br>Cases | Control<br>Cases | Total<br>number of<br>participants |
| --- | --- | --- | --- | --- | --- | --- | --- | --- |
|  |  | < 0 | Overlap 0<br>and SMD<br>< 0 | Overlap 0<br>and SMD<br>> 0 | > 0 |  |  |  |
| TRP | 106 | 48 | 48 | 8 | 2 | 5753 | 5288 | 11041 |
| TRP/CAAs | 18 | 7 | 11 | 0 | 0 | 423 | 607 | 1030 |
| CAAs | 14 | 1 | 6 | 4 | 3 | 330 | 503 | 833 |
| KYN | 67 | 19 | 30 | 15 | 3 | 4813 | 4356 | 9169 |
| KYN/TRP | 68 | 5 | 15 | 42 | 6 | 6007 | 5437 | 11444 |
| (KYN+3HK+3HA+XA+QA+PA) | 82 | 22 | 35 | 18 | 7 | 5392 | 4853 | 10245 |
| KA/KYN | 70 | 5 | 36 | 21 | 8 | 4987 | 4322 | 9309 |
| KA | 55 | 16 | 25 | 10 | 4 | 3905 | 3464 | 7369 |
| 3HK | 26 | 4 | 10 | 11 | 1 | 1364 | 1244 | 2608 |

TRP: Tryptophan, KYN: Kynurenine, KA: Kynurenic acid, 3HK: 3-Hydroxykynurenine, 3HA: 3-Hydroxyanthranilic acid, XA:
Xanthurenic acid, QA: Quinolinic acid, PA: Picolinic acid, AA: Anthranilic acid, SMD: Standardized Mean Difference, CAAs:
Competing amino acids (Valine + Phenylalanine + Tyrosine + Leucine + Isoleucine).

### Eligibility criteria

Publication in peer-reviewed journals and English language proficiency served as the key inclusion criteria for the papers in our meta-analysis. However, we also included grey literature, papers written in Thai, French, Spanish, German, Italian, and Arabic. In addition, we set inclusion criteria for observational case-control and cohort studies that evaluated the levels of TRP and TRYCATs peripherally in serum and plasma and centrally namely in CSF and brain tissues (post-mortem studies). Patients should be diagnosed in accordance with the Diagnostic and Statistical Manual of Mental Disorders (DSM) or International Classification of Diseases (ICD) criteria. Thirdly, we considered longitudinal studies that gave the baseline values of the relevant biomarkers. We excluded a) animal, genetics, and translational studies, as well as systematic reviews and meta-analyses; b) studies lacking a control group; c) studies reporting on saliva, hair, whole blood, and platelet-rich plasma samples; d) duplicate studies; and e) articles lacking the mean and standard deviation (SD) or standard error (SE) values of the measured biomarkers. Nonetheless, we asked that authors furnish us with mean (SD) values when they did not provide mean with SD or SEM values. Without more information from the authors, we estimated the mean (SD) values from the median values using the Wan et al (Wan, Wang et al. 2014) technique or computed the mean (SD) values from graphical formats using the Web Plot Digitizer (https://automeris.io/WebPlotDigitizer/).

### Primary and secondary outcomes

In the primary outcome of the current meta-analysis, we investigated the TRP and KYN levels along with IDO enzyme activity by examining the KYN/TRP ratio in affective disorders patients versus healthy control, in addition to CAAs and TRP/CAAs ratio (see **Table 1)**. Secondary outcomes involved determining the KA/KYN and 3HK/KYN ratios as indices of KAT and KMO enzymes activity respectively besides the neurotoxic TRYCAT composite (KYN+3HK+3HA+QA+XA+PA) and the solitary levels of other TRYCATs namely KA and 3HK.

### Screening and data extraction

The first two authors (AA and YT), conducted a basic evaluation of the relevant studies according to the stated inclusion criteria by examining the titles and abstracts to determine the eligibility of each research for inclusion in our meta-analysis. Then, we downloaded the complete texts of papers that met our inclusion criteria, excluding research that did meet our exclusion criteria. They used a predefined Excel file with the mean, standard deviation, and other relevant information of the included research. The final spreadsheet was double-checked by YT and AA, who contacted the third author (MM) in case of any discrepancies.

The predefined Excel file comprised of the authors’ names, the publication dates of the studies, the names and mean and standard deviation (SD) values of evaluated TRP and TRYCATs, and the sample sizes of both patient and healthy control groups. In addition, the research design, sample type (serum, plasma, CSF, and brain tissues), psychiatric assessment scales, and participants’ demographic data, including mean (SD) age, gender, and study location, were included. Furthermore, the quality of the methodology was evaluated using the immunological confounder scale (ICS) (Andrés-Rodríguez, Borràs et al. 2020). The last author modified the ICS to make it consistent with the TRYCATs research. Quality paper controls consists of two scoring scales, namely the quality and redpoints scales, which are detailed in ESF, table 2. These rating scales were largely used to evaluate the methodological quality of the publications that assessed the levels of TRYCATs in individuals with affective disorders. The score on the quality scale varied from 0 to 10, representing lower to better quality, and it focused largely on sample size, control of confounders, and sampling duration. While the redpoints scale was primarily intended to anticipate probable bias in the outcomes of TRYCATs and research designs by examining the amount of control over the important confounders. The maximum degree of control was attained when the overall score was zero; conversely, a score of twenty-six shows that the confounding variables were not considered.

**Table 2.**
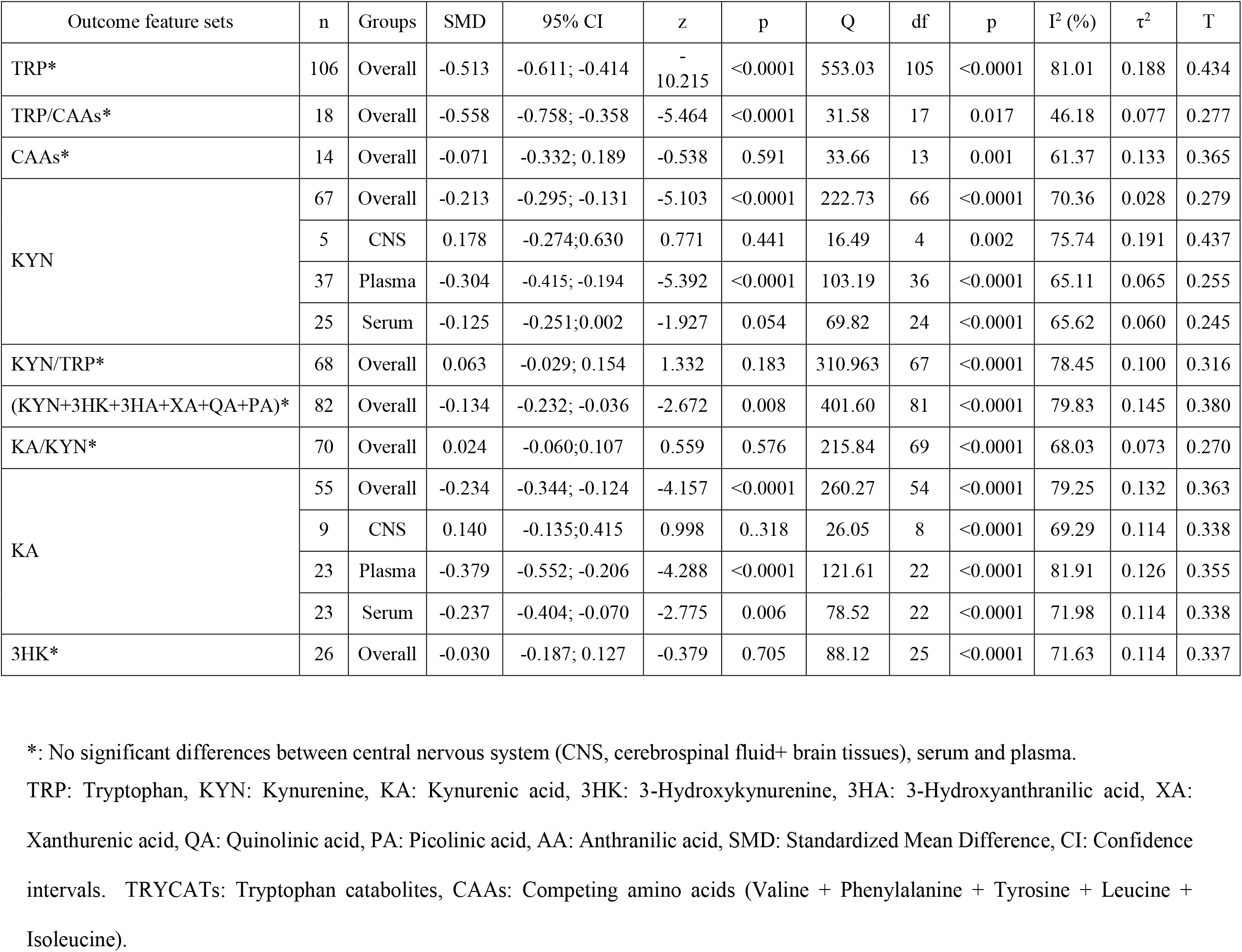
Results of meta-analysis conducted on several outcome (TRYCATs) variables with combined different media and separately.

| Outcome feature sets | n | Groups | SMD | 95% CI | z | p | Q | df | p | I<sup>2</sup> (%) | $\tau^2$ | T |
| --- | --- | --- | --- | --- | --- | --- | --- | --- | --- | --- | --- | --- |
| TRP* | 106 | Overall | -0.513 | -0.611; -0.414 | -10.215 | <0.0001 | 553.03 | 105 | <0.0001 | 81.01 | 0.188 | 0.434 |
| TRP/CAAs* | 18 | Overall | -0.558 | -0.758; -0.358 | -5.464 | <0.0001 | 31.58 | 17 | 0.017 | 46.18 | 0.077 | 0.277 |
| CAAs* | 14 | Overall | -0.071 | -0.332; 0.189 | -0.538 | 0.591 | 33.66 | 13 | 0.001 | 61.37 | 0.133 | 0.365 |
| KYN | 67 | Overall | -0.213 | -0.295; -0.131 | -5.103 | <0.0001 | 222.73 | 66 | <0.0001 | 70.36 | 0.028 | 0.279 |
|  | 5 | CNS | 0.178 | -0.274;0.630 | 0.771 | 0.441 | 16.49 | 4 | 0.002 | 75.74 | 0.191 | 0.437 |
|  | 37 | Plasma | -0.304 | -0.415; -0.194 | -5.392 | <0.0001 | 103.19 | 36 | <0.0001 | 65.11 | 0.065 | 0.255 |
|  | 25 | Serum | -0.125 | -0.251;0.002 | -1.927 | 0.054 | 69.82 | 24 | <0.0001 | 65.62 | 0.060 | 0.245 |
| KYN/TRP* | 68 | Overall | 0.063 | -0.029; 0.154 | 1.332 | 0.183 | 310.963 | 67 | <0.0001 | 78.45 | 0.100 | 0.316 |
| (KYN+3HK+3HA+XA+QA+PA)* | 82 | Overall | -0.134 | -0.232; -0.036 | -2.672 | 0.008 | 401.60 | 81 | <0.0001 | 79.83 | 0.145 | 0.380 |
| KA/KYN* | 70 | Overall | 0.024 | -0.060;0.107 | 0.559 | 0.576 | 215.84 | 69 | <0.0001 | 68.03 | 0.073 | 0.270 |
| KA | 55 | Overall | -0.234 | -0.344; -0.124 | -4.157 | <0.0001 | 260.27 | 54 | <0.0001 | 79.25 | 0.132 | 0.363 |
|  | 9 | CNS | 0.140 | -0.135;0.415 | 0.998 | 0.318 | 26.05 | 8 | <0.0001 | 69.29 | 0.114 | 0.338 |
|  | 23 | Plasma | -0.379 | -0.552; -0.206 | -4.288 | <0.0001 | 121.61 | 22 | <0.0001 | 81.91 | 0.126 | 0.355 |
|  | 23 | Serum | -0.237 | -0.404; -0.070 | -2.775 | 0.006 | 78.52 | 22 | <0.0001 | 71.98 | 0.114 | 0.338 |
| 3HK* | 26 | Overall | -0.030 | -0.187; 0.127 | -0.379 | 0.705 | 88.12 | 25 | <0.0001 | 71.63 | 0.114 | 0.337 |

\*: No significant differences between central nervous system (CNS, cerebrospinal fluid+ brain tissues), serum and plasma.
TRP: Tryptophan, KYN: Kynurenine, KA: Kynurenic acid, 3HK: 3-Hydroxykynurenine, 3HA: 3-Hydroxyanthranilic acid, XA: Xanthurenic acid, QA: Quinolinic acid, PA: Picolinic acid, AA: Anthranilic acid, SMD: Standardized Mean Difference, CI: Confidence intervals. TRYCATs: Tryptophan catabolites, CAAs: Competing amino acids (Valine + Phenylalanine + Tyrosine + Leucine + Isoleucine).

### Data analysis

The present meta-analysis was conducted using the CMA V3 program and the PRISMA criteria (ESF, table 3). To perform a meta-analysis on TRP or a particular TRYCAT, at least three studies on that TRYCAT were necessary. The neurotoxicity index and ratios were compared between patients and controls by computing the mean values of the outcomes while assuming dependence. Thus, we compared the neurotoxic TRYCATs profile of patients with affective disorders and healthy controls by entering the relevant TRYCATs in the meta-analysis, and we analyzed the KYN/TRP, KA/KYN and 3HK/KYN ratios as an indexes for IDO, KAT and KMO enzyme activities respectively (Almulla, Vasupanrajit et al. 2022). IDO enzyme activity is estimated by selecting a positive effect size direction for increasing KYN and a negative direction for lowered TRP levels in patients, KAT activity was assessed by setting KA as positive and KYN as negative, KMO by setting 3HK as positive and KYN as negative, and the TRP/CAAs ratio by setting TRP as positive and CAA as negative. We utilized the random-effects model with constrained maximum likelihood to pool the effect sizes since participants characteristics were not homogeneous across trials in the current meta-analysis. We report the effect size as the standardized mean difference (SMD) with 95 percent confidence intervals (95% CI), and statistical significance was defined as a two-tailed p-value less than 0.05. The effect size was described as large, moderate, and small based on the SMD values of 0.80, 0.5, and 0.20, respectively (Cohen 1988). In accordance with prior meta-analyses (Almulla, Vasupanrajit et al. 2022, Vasupanrajit, Jirakran et al. 2022), heterogeneity was determined by calculating tau-squared statistics, although we also show the Q and I^2^ metrics. To identify the causes of heterogeneity in the present meta-analysis, a meta-regression was conducted. We used subgroup analysis to identify differences in TRP and TRYCATs in different media, including in CNS (brain tissues + CSF), serum and plasma, as well as the distinctions between MDD and BD. Each of the above categories serves as a unit of analysis. Since there are no indications of statistically significant differences between studies that measured CSF and brain tissues in the biomarkers, we merged the CSF and brain tissues under the umbrella name CNS. We show the results in the combined study group of affective disorders (MDD + BD), and when there are significant differences, we show the results in MDD and BD, separately. We conducted sensitivity analyses using the leave-one-out approach to examine the robustness of the impact sizes and heterogeneity across studies. Publication bias was investigated using the fail-safe N technique, continuity-corrected Kendall tau, and Egger’s regression intercept, using one-tailed p-values for the last two approaches. In the face of an asymmetry shown by Egger’s test, we used the trim-and-fill approach developed by Duval and Tweedie to impute the missing studies and compute adjusted effect sizes. To discover small study effects, we also used funnel plots (study precision vs SMD), which concurrently show observed and imputed missing values.

**Table 3.**
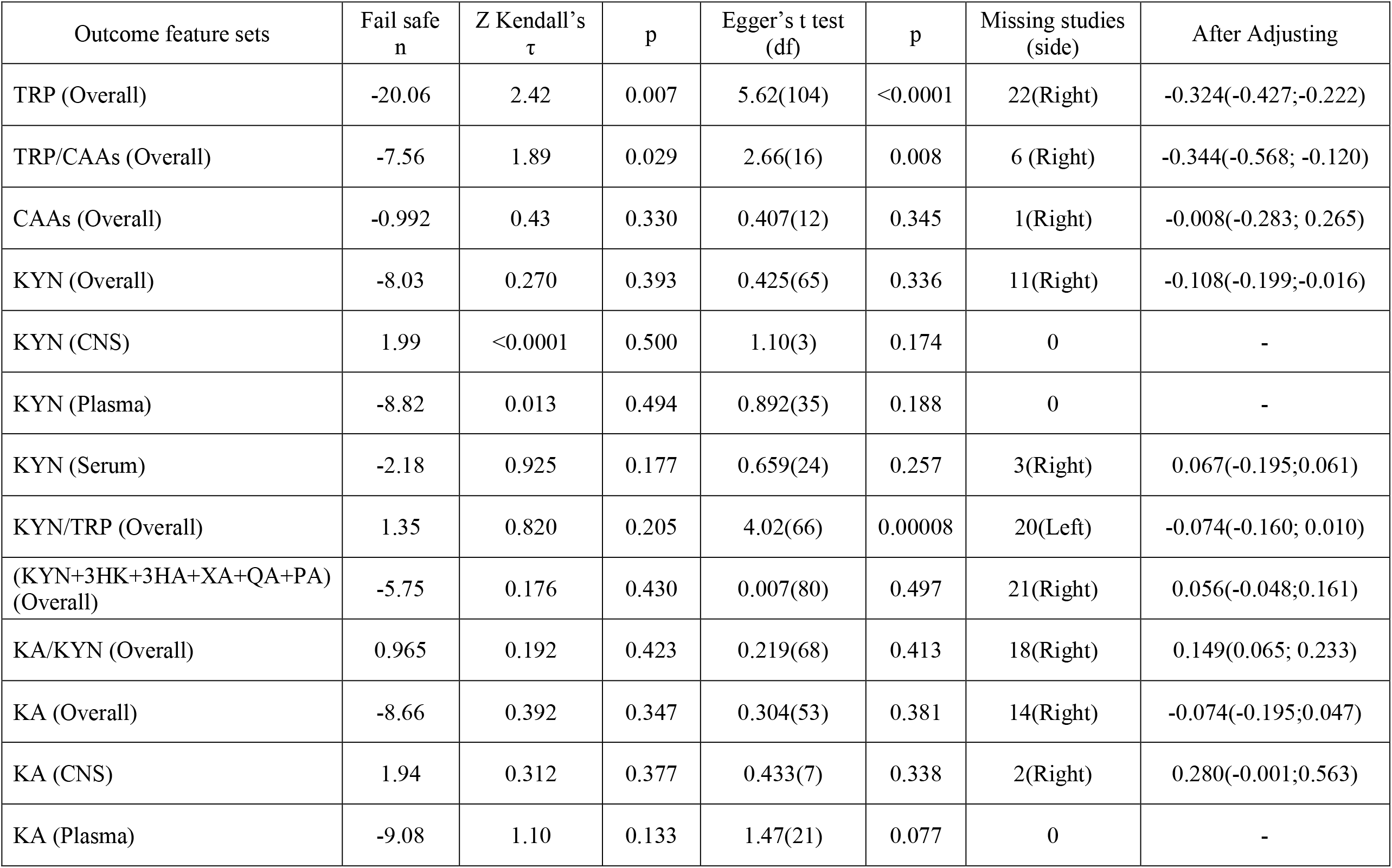
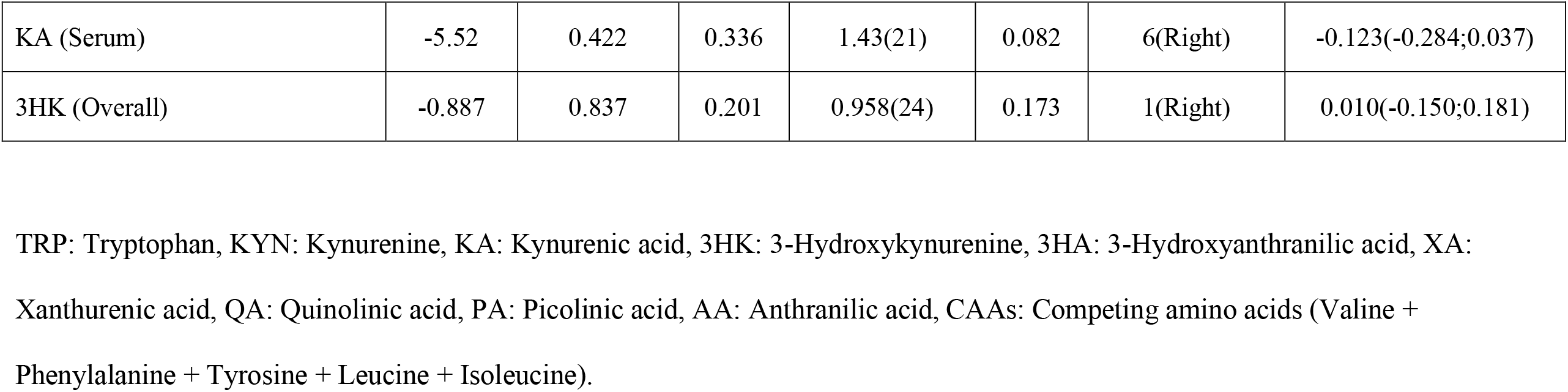
Results on publication bias.

| Outcome feature sets | Fail safe<br>n | Z Kendall's<br>$\tau$ | p | Egger's t test<br>(df) | p | Missing studies<br>(side) | After Adjusting |
| --- | --- | --- | --- | --- | --- | --- | --- |
| TRP (Overall) | -20.06 | 2.42 | 0.007 | 5.62(104) | <0.0001 | 22(Right) | -0.324(-0.427;-0.222) |
| TRP/CAAs (Overall) | -7.56 | 1.89 | 0.029 | 2.66(16) | 0.008 | 6 (Right) | -0.344(-0.568; -0.120) |
| CAAs (Overall) | -0.992 | 0.43 | 0.330 | 0.407(12) | 0.345 | 1(Right) | -0.008(-0.283; 0.265) |
| KYN (Overall) | -8.03 | 0.270 | 0.393 | 0.425(65) | 0.336 | 11(Right) | -0.108(-0.199;-0.016) |
| KYN (CNS) | 1.99 | <0.0001 | 0.500 | 1.10(3) | 0.174 | 0 | - |
| KYN (Plasma) | -8.82 | 0.013 | 0.494 | 0.892(35) | 0.188 | 0 | - |
| KYN (Serum) | -2.18 | 0.925 | 0.177 | 0.659(24) | 0.257 | 3(Right) | 0.067(-0.195;0.061) |
| KYN/TRP (Overall) | 1.35 | 0.820 | 0.205 | 4.02(66) | 0.00008 | 20(Left) | -0.074(-0.160; 0.010) |
| (KYN+3HK+3HA+XA+QA+PA)<br>(Overall) | -5.75 | 0.176 | 0.430 | 0.007(80) | 0.497 | 21(Right) | 0.056(-0.048;0.161) |
| KA/KYN (Overall) | 0.965 | 0.192 | 0.423 | 0.219(68) | 0.413 | 18(Right) | 0.149(0.065; 0.233) |
| KA (Overall) | -8.66 | 0.392 | 0.347 | 0.304(53) | 0.381 | 14(Right) | -0.074(-0.195;0.047) |
| KA (CNS) | 1.94 | 0.312 | 0.377 | 0.433(7) | 0.338 | 2(Right) | 0.280(-0.001;0.563) |
| KA (Plasma) | -9.08 | 1.10 | 0.133 | 1.47(21) | 0.077 | 0 | - |
| KA (Serum) | -5.52 | 0.422 | 0.336 | 1.43(21) | 0.082 | 6(Right) | -0.123(-0.284;0.037) |
| 3HK (Overall) | -0.887 | 0.837 | 0.201 | 0.958(24) | 0.173 | 1(Right) | 0.010(-0.150;0.181) |

TRP: Tryptophan, KYN: Kynurenine, KA: Kynurenic acid, 3HK: 3-Hydroxykynurenine, 3HA: 3-Hydroxyanthranilic acid, XA:
Xanthurenic acid, QA: Quinolinic acid, PA: Picolinic acid, AA: Anthranilic acid, CAAs: Competing amino acids (Valine +
Phenylalanine + Tyrosine + Leucine + Isoleucine).

## Results

### Search results

Figure 2. shows the PRISMA flow chart with the overall search outcomes and the number of included and omitted articles. We investigated 11038 articles of the initial searching processes, which relied on our specific keywords and mesh terms (as listed in ESF, table 1). However, we refined the search’s results and eliminated 10172 duplicate and irrelevant studies. After applying our inclusion-exclusion criteria, 124 full-text eligible articles were included in the current systematic review. Due to exclusion criteria mentioned in ESF, table 4, three out of these 124 studies were excluded. Hence, the current meta-analysis involved 121 studies (Coppen, Brooksbank et al. 1972, Coppen, Eccleston et al. 1973, Moller, Kirk et al. 1976, Wood, Harwood et al. 1978, Shaw, Tidmarsh et al. 1980, DeMyer, Shea et al. 1981, Healy, Carney et al. 1982, Møller, Kirk et al. 1982, Menna-Perper 1983, Gerner, Fairbanks et al. 1984, Joseph, Brewerton et al. 1984, Guicheney, Léger et al. 1988, Cowen, Parry-Billings et al. 1989, Anderson, Parry-Billings et al. 1990, Chiaroni, Azorin et al. 1990, Maes, Jacobs et al. 1990, Russ, Ackerman et al. 1990, Price, Charney et al. 1991, Lucca, Lucini et al. 1992, Quintana 1992, Maes, Meltzer et al. 1993, Møller 1993, Ortiz, Mariscot et al. 1993, Karege, Widmer et al. 1994, Maes, De Backer et al. 1995, Maes, Wauters et al. 1996, Maes, Verkerk et al. 1997, Mauri, Ferrara et al. 1998, Song, Lin et al. 1998, Moreno, Gelenberg et al. 1999, Hoekstra, van den Broek et al. 2001, Mauri, Boscati et al. 2001, Porter, Gallagher et al. 2003, Rief, Pilger et al. 2004, Hayward, Goodwin et al. 2005, Hoekstra, Fekkes et al. 2006, Miller, Llenos et al. 2006, Myint, Kim et al. 2007, Myint, Kim et al. 2007, Czermak, Hauger et al. 2008, Manjarrez-Gutierrez, Marquez et al. 2009, Roiser, Levy et al. 2009, Gabbay, Klein et al. 2010, Olsson, Samuelsson et al. 2010, Maes, Galecki et al. 2011, Steiner, Walter et al. 2011, Sublette, Galfalvy et al. 2011, Ebesunun, Eruvulobi et al. 2012, Hughes, Carballedo et al. 2012, Maes and Rief 2012, Pinto, de Souza et al. 2012, Xu, Fang et al. 2012, Erhardt, Lim et al. 2013, Hennings, Schwarz et al. 2013, Moreno, Gaspar et al. 2013, Quak, Doornbos et al. 2014, Reininghaus, McIntyre et al. 2014, Bay-Richter, Linderholm et al. 2015, Bradley, Case et al. 2015, Busse, Busse et al. 2015, Dahl, Andreassen et al. 2015, Hüfner, Oberguggenberger et al. 2015, Nikkheslat, Zunszain et al. 2015, Savitz, Dantzer et al. 2015, Savitz, Dantzer et al. 2015, Savitz, Drevets et al. 2015, Savitz, Drevets et al. 2015, Teraishi, Hori et al. 2015, Brundin, Sellgren et al. 2016, Clark, Pocivavsek et al. 2016, Georgin-Lavialle, Moura et al. 2016, Meier, Drevets et al. 2016, Schwieler, Samuelsson et al. 2016, Veen, Myint et al. 2016, Yoshimi, Futamura et al. 2016, Young, Drevets et al. 2016, Baranyi, Amouzadeh-Ghadikolai et al. 2017, Birner, Platzer et al. 2017, Cho, Savitz et al. 2017, Hu, Li et al. 2017, Krause, Myint et al. 2017, Platzer, Dalkner et al. 2017, Sorgdrager, Doornbos et al. 2017, Umehara, Numata et al. 2017, Wurfel, Drevets et al. 2017, DeWitt, Bradley et al. 2018, Doolin, Allers et al. 2018, Kuwano, Kato et al. 2018, Liu, Ding et al. 2018, Moaddel, Shardell et al. 2018, Mukherjee, Krishnamurthy et al. 2018, Ogawa, Koga et al. 2018, Pan, Xia et al. 2018, Poletti, Myint et al. 2018, Wu, Mai et al. 2018, Wu, Zhong et al. 2018, Zhou, Zheng et al. 2018, Aarsland, Leskauskaite et al. 2019, Castillo, Murata et al. 2019, Heilman, Hill et al. 2019, Krause, Kirnich et al. 2019, Poletti, Melloni et al. 2019, Pompili, Lionetto et al. 2019, Sellgren, Gracias et al. 2019, Zhou, Zheng et al. 2019, Achtyes, Keaton et al. 2020, Carrillo-Mora, Pérez-De la Cruz et al. 2020, Colle, Masson et al. 2020, Erabi, Okada et al. 2020, Murata, Murphy et al. 2020, Ryan, Allers et al. 2020, Sakurai, Yamamoto et al. 2020, Steen, Dieset et al. 2020, Sun, Drevets et al. 2020, van den Ameele, van Nuijs et al. 2020, Cathomas, Guetter et al. 2021, Chiu, Yang et al. 2021, Milaneschi, Allers et al. 2021, Öztürk, Yalın Sapmaz et al. 2021, Trepci, Sellgren et al. 2021, Paul, Schwieler et al. 2022).

**Figure 2:**
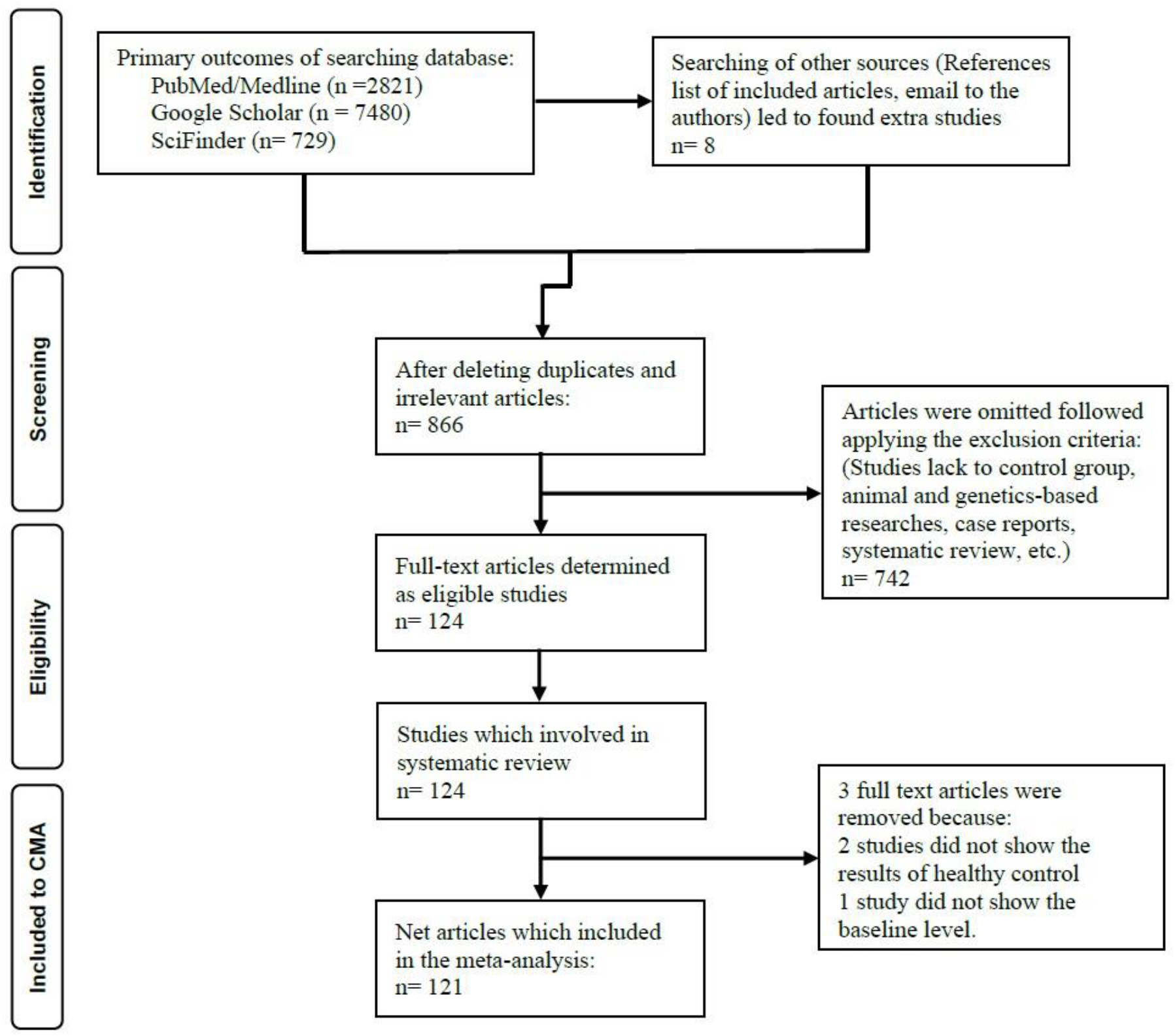
The PRISMA flow chart.

Ten of the eligible articles examined the TRP and TRYCATs in MDD and BD within the same study. Three studies assessed centrally and peripherally levels of TRP and TRYCTAs. Furthermore, 1 study involved two separate cohorts of patients and healthy controls. Hence, in the present meta-analysis, the overall effect size was pooled from 135 (16 CNS, 84 plasma, 35 serum) studies, namely 110 studies (10 CNS, 71 plasma, 29 serum) in MDD patients and 25 studies (6 CNS, 13 plasma, 6 serum) in BD patients. The total recruited number of individuals was 15470 in the present meta-analysis, distributed as 8024 patients with affective disorders and 7446 control subjects. The age of the participants extended from 16 to 69 years.

As shown in ESF, table 5, our systematic review found that high performance liquid chromatography (HPLC) was the most often used method for measuring TRP and TRYCATs, as it has been deployed in 50 papers. Numerous nations contributed to the included studies; the United States contributed the most with 29 studies, followed by the United Kingdom with 10 studies; the remainder of the contributors were as follows: 9 from Sweden, 8 are from China and Germany, 7 are from Belgium, 6 are from Japan, the Netherlands, and Italy, 5 are from Austria, 4 are from Ireland, 3 are from Denmark, France, Mexico and Switzerland 2 are from Norway, South Korea and Spain, and only one is from Taiwan, Turkey, Brazil, Canada, and Nigeria. We analyzed the quality and redpoints whose median (min-max) ratings were 5 (min=1, max=8,75), 13,5 (min=4,5, max=23), respectively, and the results are presented in table 5 of ESF.

### Primary outcome variables

#### TRP and the TRP/CAAs ratio

The present meta-analysis identified TRP measurements in 106 research papers. Table 1 shows that in 48 studies the CI were entirely negative of zero, whereas there were only 2 studies which showed CI intervals that were entirely positive of zero. There were 56 studies that showed CI overlapping with zero and of those 48 showed SMD values smaller than zero, and 8 papers showed SMD values greater than zero. In individuals with affective disorders, TRP was significantly decreased with a modest effect size of -0.513 as shown in **Table 2**. There was some publication bias as shown in **Table 3** with 22 papers missing on the right side of the funnel plot. Nevertheless, after imputing the missing studies, the adjusted point estimate remained significant at SMD=-0.324.

We conducted a subgroup analysis to investigate the differences between total and free TRP levels in patients versus controls. The results indicated no significant difference (p=0.176) between total and free TRP levels, whilst both were decreased in affective disorders patients.

Table 2 shows that the TRP/CAAs ratio was significantly lower in patients than in controls with a modest effect size of -0.558. Table 3 shows a bias with 6 studies missing on the right side of the funnel plot and imputing these studies lowered the SMD value although it remained significant. CAAs results were obtained from 14 studies. Tables 1 and 2 show that CAAs levels were not significantly different between patients and controls.

#### KYN and the KYN/TRP ratio

We included 67 (5 CNS, 37 plasma and 29 serum) KYN studies in the present systematic review. Table 1 shows the CI distributions of the KYN and KYN/TRP data. Table 2 shows that KYN was significantly lower in patients than in controls with a small effect size. Nevertheless, group analysis showed a significant difference (p=0.025) between CNS, plasma, and serum and that KYN levels were significantly reduced in plasma albeit with a modest impact size, but not in the CNS or serum. Egger’s regression and Kendall’s tau revealed publishing bias in serum and imputing 3 studies to the right reduced the SMD value to 0.067, whereas there was no bias in the CNS and plasma studies (see table 3). No significant difference was found in the KYN/TRP ratio between patients and controls as shown in table 2.

### Secondary outcome variables

The effect size of the neurotoxicity composite (KYN+3HK+3HA+PA+XA+QA) was obtained from 82 studies. Table 2 shows that patients with affective disorders have a significantly reduced neurotoxicity composite index, albeit with a small effect size (SMD=-0.134). There was evidence of bias with 21 studies on the right and adjusting the effect size for these studies changed the results to non-significant (see Table 3). Table 2 shows no significant difference in the KA/KYN ratio between patients and controls, although after imputing 18 missing studies on the right site, the KA/KYN ratio was increased (see Table 3). There were significant differences in KA results between CNS, plasma and serum with plasma and serum KA being significantly decreased in patients. Nevertheless, after imputing missing studies the differences in serum KA were no longer significant. Table 2 shows no significant differences in 3HK between the groups.

### Meta-regression analyses

We conducted meta-regression analysis to identify the most likely causes of the heterogeneity in TRP and TRYCAT data. As demonstrated in ESF, table 6, the unmedicated status affected TRP and CAAs. In addition, TRP was impacted by male gender and sample size. Age and severity are likely other factors that impact part of the variability. The higher number of studies compared to prior meta-analyses influenced all outcomes in ESF table 6 except 3HK, CAAs, and TRP/CAAs ratio.

## Discussion

### 1. Tryptophan availability to the brain

The first major finding of this large-scaled systematic review is that total and free TRP and the TRP/CAAs ratio were significantly lower with a moderate effect size in MDD and BD patients than in controls and that there were no differences between MDD and BD. The current findings are consistent with the results of prior meta-analyses (Ogawa, Fujii et al. 2014, Marx, McGuinness et al. 2021), although the meta-analysis conducted by Arnone et al. found unaltered TRP in MDD and BD patients (Arnone, Saraykar et al. 2018). The amount of TRP which will reach the brain depends not only on the concentrations of peripheral, total and free TRP, but also on CAAs and the TRP/CAAs ratio (Yuwiler, Oldendorf et al. 1977, Pardridge 1979). Brain TRP concentrations are influenced by CAAs since the latter are competing with TRP for transport through the large amino acid transporter 1 (LAT 1) of the blood brain barrier (BBB) (Fernstrom, Larin et al. 1973, Pardridge 1979). In this respect, our study found that the CAAs levels were unaltered in MDD/BD indicating that the lowered TRP/CAAs ratio is explained by lowered TRP. Such findings were indeed reported in previous studies (DeMyer, Shea et al. 1981, Maes, Meltzer et al. 1993, Maes, Wauters et al. 1996, Song, Lin et al. 1998).

### 2. KYN and IDO

The second major finding of the current research is that peripheral KYN levels were significantly lower in patients than in controls, whereas the KYN/TRP ratio was unaltered. The present KYN findings are consistent with preceding meta-analyses (Ogyu, Kubo et al. 2018, Bartoli, Misiak et al. 2021, Hebbrecht, Skorobogatov et al. 2021, Marx, McGuinness et al. 2021), although Arnone et al. reported normal KYN levels in BD patients (Arnone, Saraykar et al. 2018). Nevertheless, previous studies were performed on a smaller number of studies and did not consider possible differences among central, serum and plasma KYN assays. In this respect, subgroup analysis revealed that peripheral KYN levels were lowered in the plasma of patients but not in their serum. All in all, since KYN is not altered in serum and CNS and since plasma KYN levels are more difficult to interpret (see below), the KYN data indicate decreased or unchanged levels in the major affective mood disorders.

### 3. TRYCAT neurotoxicity in affective disorders

A third major finding of the current systematic review is that patients with affective disorders showed no significant changes in the neurotoxicity index (after correcting for possible bias) comprising KYN, 3HK, 3HA, PA, XA and QA. Moreover, patients showed a significant increase in the KA/KYN ratio (an indicator of lowered KYN-associated neurotoxicity) and a reduction in KA in peripheral blood only. Previous meta-analysis also reported a decrease in peripheral KA levels, although they did not measure CNS KA levels (Ogyu, Kubo et al. 2018, Bartoli, Misiak et al. 2021, Hebbrecht, Skorobogatov et al. 2021, Marx, McGuinness et al. 2021). The current findings suggest that the IDO enzyme and production of neurotoxic TRYCATs are not upregulated in patients with mood disorders, which partially contradicts the IDO theory of affective disorders, postulating that IDO stimulation with decreased TRP and elevated neurotoxic TRYCATs is involved in the pathophysiology of MDD/BD (Maes, Leonard et al. 2011).

### 4. Heterogeneity

In the current meta-analysis, we found a high degree of heterogeneity in the TRP and TRYCATs data, and our meta-regression and group analyses uncovered significant sources of heterogeneity. Discrepancies between measurements in CNS, plasma and serum KYN were also detected in a previous meta-analysis in schizophrenia (Almulla, Vasupanrajit et al. 2022). In that paper, we have discussed that plasma is not the most accurate medium for determining KYN (and other TRYCATs) because plasma assays are more susceptible to pre-analytical and analytical errors (Almulla, Vasupanrajit et al. 2022) resulting from a) probable degradation of KYN and TRP in response to the presence of carbonyl-containing compounds, namely EDTA (Bellmaine, Schnellbaecher et al. 2020), b) dilutional effects (particularly on small amounts of analytes) of anticoagulants in plasma tubes (Sotelo-Orozco, Chen et al. 2021), and c) increased α-amino products due to EDTA decomposition in high temperatures (Parvy, Bardet et al. 1983). Moreover, previous studies recommended serum to assay TRP since the anticoagulants within plasma tubes may contaminate the measurements, particularly when using HPLC and spectrophotometers techniques (Davidson 2002, Kulkarni, Karanam et al. 2016). In addition to the impact of differences between serum, plasma, and CNS, other important sources of heterogeneity include a) the medicated status of the patients, with unmedicated patients exhibiting larger effect sizes in most biomarkers compared to treated patients, and b) sex which impacts the KYN/TRP ratio, and c) to a lesser extent, age and sample size (see ESF, table 6).

### 5. Interpretation of the results

The IDO-neurotoxicity theory of affective disorders was developed in response to the observation that IFN-α-based immunotherapy causes depression and that increased IRS responses and production of neurotoxic TRYCAT levels are directly related to the onset of this type of depression (Bonaccorso, Marino et al. 2002). Nonetheless, our negative results concerning TRYCAT and IDO levels do not support the theory that activation of IDO/TDO is involved in MDD/BD (Maes 2015). While the TRYCAT pathway is activated during severe IRS responses, such as acute COVID-19 infection (Almulla, Supasitthumrong et al. 2022) and IFN-α therapy (Bonaccorso, Marino et al. 2002), no such changes may be observed in conditions of mild chronic inflammation, such as MDD/BD (this study) and Alzheimer’s disease (Almulla, Supasitthumrong et al. 2022). Since plasma/serum TRP concentrations are around 50 μmol/L while KYN concentrations are around 3 μmol/L, TRP is abundantly available as a substrate for KYN formation. Therefore, it is plausible that IDO (TDO) may be inhibited in MDD/BD rather than that lowered substrate availability determines the downregulation of the pathway. First, since the substrate TRP is reduced in MDD/BD patients, the IDO enzyme may be self-regulating and transformed to an inactive state (Nelp Micah, Kates Patrick et al. 2018). When TRP concentrations are low, catalytically inactive ferric IDO1 may accumulate during turnover and the enzyme may autooxidize (Booth, Basran et al. 2015). Second, the IDO enzyme is inhibited by nitric oxide (NO), which is significantly increased in MDD and BD (Savaş, Herken et al. 2002, Talarowska, Gałecki et al. 2012, Maes, Simeonova et al. 2019).

All in all, our results show that mood disorders are characterized by lowered TRP levels while IDO and the TRYCAT pathway are not stimulated. The majority of TRP in the circulation is bound to albumin (the total TRP pool) and, therefore, any changes in serum albumin will impact total TRP levels (Mc and Oncley 1958, Fernstrom, Larin et al. 1973). Some studies found that serum albumin levels were lower in people with depression and that there was a link between serum albumin and total serum/plasma TRP (Maes, Vandewoude et al. 1991, Maes, Wauters et al. 1996, Maes, Smith et al. 1997, Liu, Zhong et al. 2015). As such, reduced albumin levels, due to the acute phase or mild chronic IRS response in affective disorders, may predispose towards lowered serotonin synthesis in the brain (Maes, Vandewoude et al. 1991, Maes, Wauters et al. 1996, Maes, Smith et al. 1997). All in all, the lowered TRP concentrations are at least in part the consequence of IRS activation in MDD/BD and, in fact, constitute a CIRS response aimed to attenuate hyperinflammation and combat infections (Maes, Leonard et al. 2011). Other mechanisms for decreased TRP availability include a) platelets being activated in MDD (Moreno, Gaspar et al. 2013), which may be accompanied by an increased TRP uptake; and b) the circulatory levels of free fatty acids, which are in part mediated by insulin levels (Almulla, Vasupanrajit et al. 2022). Because serum/plasma TRP availability influences TRP concentrations in the brain (Fernstrom, Larin et al. 1973, Curzon 1979), decreased TRP concentrations in peripheral blood may influence serotonin synthesis in the brain, which is thought to play a role in MDD/BD (Coppen 1967, Lapin and Oxenkrug 1969, Maes and Meltzer 1995, Oxenkrug 2013).

The lowered levels of TRP may suggest that MDD/BD patients show lowered neuroprotection. First, TRP and serotonin are antioxidants (Xu, Liu et al. 2018), and serotonin has neuroprotective properties by preserving neuroplasticity and preventing neuronal injuries (Croonenberghs, Verkerk et al. 2005, Radulescu, Dragoi et al. 2021). In fact, the antioxidant role of TRP is indirect (Perez-Gonzalez, Muñoz-Rugeles et al. 2014, Pérez-González, Alvarez-Idaboy et al. 2015) via metabolites such as melatonin, serotonin, 3-HK and XA (Reiter, Tan et al. 1999). Second, low levels of some TRYCATs, including KYN, KA, XA and QA, may negatively impact neuroprotection because these TRYCATs have anti-inflammatory effects for example by lowering the IFN-γ/IL-10 ratio (Maes, Mihaylova et al. 2007), whilst KA, 3HK, 3HA, and XA have antioxidant effects (Goda, Hamane et al. 1999, Maes, Leonard et al. 2011). In addition, KA has a neuroprotective role by downregulating the excitatory receptors in the brain, namely, NMDA, α-amino-3-hydroxy-5-methyl-4-isoxazolepropionic acid (AMPA), and kainate glutamate ionotropic receptors and by impeding the alpha 7 nicotinic acetylcholine receptor (Morris, Carvalho et al. 2016).

### Limitations

This study has shown a number of limitations that should be considered in future investigations. First, few studies reported on the central levels of TRP and TRYCATs and, therefore, more research should focus on postmortem tissue and CSF concentrations of TRP and TRYCATs. Second, a substantial proportion of the studies included in our analysis lacked information about the treatment histories of patients. This may be relevant because at least some antidepressants, such as escitalopram, are associated with a lower plasma KYN/TRP ratio due to a decrease in KYN but not TRP (Sun, Drevets et al. 2020). Thus, it is preferable to thoroughly investigate the TRP and TRYCATs in drug-naïve patients with a first episode or to control statistically for the medication status. Moreover, future research should stratify patients according to the staging phases of affective disorders since disease staging is related to greater neurotoxicity (Maes, Moraes et al. 2019, Maes, Rachayon et al. 2022). Fourth, a considerable number of studies seemed to have been conducted without correcting for major confounding variables such as smoking and alcoholism and their effects on TRP (Badawy 2002) and TRYCAT (Leclercq, Schwarz et al. 2021) metabolism. Importantly, it may be that IDO activation is a hallmark of somatization rather than of affective disorders (Maes, Galecki et al. 2011) and, therefore, future research in affective disorders should always include measurement of somatization and examine the comorbidity between MDD/BD and somatization. It may be that the assay of IgA responses to TRYCAT adducts is much more sensitive than the methods used in the studies included in our meta-analysis. Thus, while the neurotoxic TRYCATs assessed with the conventional methods were not associated with schizophrenia (Almulla, Vasupanrajit et al. 2022), we found that IgA responses to neurotoxic TRYCATs were strongly associated with the severity of the phenome of schizophrenia(Kanchanatawan, Sirivichayakul et al. 2018).

The majority, if not all, studies compared TRP and TRYCATs between patients diagnosed with a major depressive episode per DSM or ICD criteria and controls. Recently, however, we have demonstrated that such diagnostic criteria are grossly inadequate (Maes and Stoyanov 2022) and that precision nomothetic psychiatry has enabled the construction of an endophenotype class of severely depressed patients (termed Major Dysmood Disorder) with immune-inflammatory and nitro-oxidative disorders (Maes, Moraes et al. 2019, Maes, Rachayon et al. 2022, Maes and Stoyanov 2022). This is the target class for detecting IDO-induced changes in TRYCATs.

## Conclusion

The key findings of the present systematic review and meta-analysis are summarized in Figure 1. Both MDD and BD are associated with central and peripheral TRP depletion, which may be explained by lowered serum albumin levels. IDO enzyme activity did not exhibit signs of hyperactivity, as indicated by the patients’ lowered KYN levels and an unchanged KYN/TRP ratio. Moreover, there is no evidence that MDD and BD are accompanied by increased neurotoxicity due to an activated TRYCAT pathway. Future research should employ the precision nomothetic approach and control for the use of antidepressants (Maes and Stoyanov 2022) to delineate the involvement of serum and CNS (not plasma) TRP and TRYCATs in Major Dysmood Disorder and not a major depressive episode according to DSM/ICD criteria.

## Data Availability

The last author (MM) will respond to any reasonable request for the dataset (Excel file) employed in the current meta-analysis after it have been fully exploited by all authors.

## Ethical approval and consent to participate

Not applicable.

## Consent for publication

Not applicable.

## Author’s contributions

The study was designed by AA and MM. AA, YT, and AV collected the data. AA and MM performed the statistical analysis. All authors contribute to the writing of the paper and approved submission of the final draft.

## Acknowledgments

Not applicable.

