## supplementary file for "The tryptophan catabolite or kynurenine pathway in major depressive and bipolar disorder: a systematic review and meta-analysis"

SHORT TITLE: Kynurenine pathway in affective disorders

Abbas F. Almulla, Ph.D.[a,b] , Yanin Thipakorn, M.D., Ph.D.[a], Asara Vasupanrajit, MS.c[a], Ali Abbas Abo Algon, MS.c [c], Chavit Tunvirachaisakul, M.D., Ph.D.[a] Ashwan Abdulzahra Hashim Aljanabi M.D., Ph.D [d,e]., Gregory Oxenkrug, M.D., Ph.D.[f], Hussein K. Al-Hakeim, Ph.D.[g], Michael Maes, M.D., Ph.D.[a,h,i]

[a] Department of Psychiatry, Faculty of Medicine, Chulalongkorn University, Bangkok, Thailand.

[b] Medical Laboratory Technology Department, College of Medical Technology, The Islamic University, Najaf, Iraq.

[c] Iraqi Education Ministry- Najaf- Iraq.

[d] Department of Medicine, Faculty of Medicine, University of Kufa,

[e] Department of Psychiatry Al-Hakeem General Hospital

[f] Department of Psychiatry, Tufts University School of Medicine and Tufts Medical Center, Boston, MA 02111, USA.

[g] Department of Chemistry, College of Science, University of Kufa, Kufa, Iraq.

[h] Department of Psychiatry, Medical University of Plovdiv, Plovdiv, Bulgaria.

[i] Department of Psychiatry, IMPACT Strategic Research Centre, Deakin University, Geelong, Victoria, Australia.

**Corresponding author:**

Prof. Dr Michael Maes, M.D., Ph.D.

Department of Psychiatry

Faculty of Medicine, Chulalongkorn University

Bangkok, 10330

Thailand

E-mail addresses:

**ESF, Table 1.** Search sentences and terms were used in each database

| Database Name | Search Sentence | No. of Articles |
|---|---|---|
| PubMed/Medline | (((((((((((TRYCATs and MDD) OR (TRYCATs and MDD)) OR (Tryptophan Catabolites and MDD)) OR (L-TRP and MDD)) OR (L-KYN and MDD)) OR (KYNA and MDD)) OR (ANA and MDD)) OR (3-HK and MDD)) OR (XA and MDD)) OR (3-HA and MDD)) OR (QA and MDD)) OR (PA and MDD) | **2133** |
| | ((((((((((Bipolar and TRYCATs)* OR (Bipolar and Tryptophan)) OR (Bipolar and tryptophan cata)) OR (Bipolar and TRYCATs)) OR (Bipolar and TRP)) OR (BD and KYN)) OR (BD and kynurenine)) OR (Bipolar Kynurenine pathway)) OR (BD and kynurenine pathway)) OR (Bipolar and IDO)) OR (Bipolar and TDO) | **506** |
| | ((((((((((Unipolar and TRYCATs)* OR (Unipolar and Tryptophan)) OR (Unipolar and tryptophan cata)) OR (Unipolar and TRYCATs)) OR (Unipolar and TRP)) OR (UD and KYN)) OR (UD and kynurenine)) OR (Unipolar Kynurenine pathway)) OR (UD and kynurenine pathway)) OR (Unipolar and IDO)) OR (Unipolar and TDO) | **105** |
| | ((((((((((Mania and TRYCATs)* OR (Mania and Tryptophan)) OR (Hypomania and tryptophan cata)) OR (Hypomania and TRYCATs)) OR (Mania and TRP)) OR (Mania and KYN)) OR (Mania and kynurenine)) OR (Mania Kynurenine pathway)) OR (Mania and kynurenine pathway)) OR (Mania and IDO)) OR (Mania and TDO | **77** |
| Google Scholar | (((((Depression* and IDO activation) OR (TDO activation)) OR (Kynurenine pathway)) OR (KMO activation)) OR (Tryptophan degradation)) AND (Decreased Tryptophan) | **4960** |
| | (((((Bipolar Disorder* and IDO activation) OR (TDO activation)) OR (Kynurenine pathway)) OR (KMO activation)) OR (Tryptophan degradation)) AND (Decreased Tryptophan) | **1750** |
| | (((((Mania* and IDO activation) OR (TDO activation)) OR (Kynurenine pathway)) OR (KMO activation)) OR (Tryptophan degradation)) AND (Decreased Tryptophan) | **770** |

| | | |
|---|---|---|
| **SciFinder** | Major depressive disorder and kynurenine pathway, Major depressive disorder and tryptophan catabolism pathway, Major depressive disorder and TRYCATs, Major depressive disorder and kynurenine, Major depressive disorder and tryptophan, Major depressive disorder kynurenic acid, Depression and kynurenine pathway | **668** |
| | Bipolar Disorder and kynurenine pathway, Bipolar Disorder and tryptophan catabolism pathway, Bipolar Disorder and TRYCATs, Bipolar Disorder and kynurenine, Bipolar Disorder and tryptophan, Bipolar Disorder kynurenic acid, Bipolar Disorder and kynurenine pathway | **61** |

**ESF, Table 2.** Immune cofounder's scale (ICS) applied from Andrés-Rodríguez, et al., 2019

| **Methodological quality of the study** | | |
|---|---|---|
| **1** | Study sample ≥ 128 participants including patients and controls (1= Yes, 0 = No) | |
| **2** | Did the study control the results for potential confounders (e.g., age, BMI, gender, race)? (1= Yes, 0 = No) | |
| **3** | Were participants with affective disorders and controls age- and-gender-matched or was there a statistical control? (1= Yes, 0 = No) | |
| **4** | Was the time of sample collection specified (e.g., morning vs. evening)? (1= Yes, 0 = No) | |
| **5** | Were participants with affective disorders free of immunomodulatory drugs including anti-cytokines, glucocorticoids, immunoglobulins, and immunosuppressants, or was there a medication washout period, or was drug intake statistically controlled for? (1= Yes, 0 = No) | |
| **6** | Were participants with affective disorders free of antidepressants and mood stabilizers or were the data statistically controlled for? (1= Yes, 0 = No) | |
| **7** | Reporting either the manufacturer of the test or detection limit and coefficients of variation (1= Yes, 0 = No) | |
| **8** | Reporting how data under detection limit were handled (1 = Yes, 0 = No) | |
| **9** | Reporting % of the sample under detection limit (1=Yes, 0= No) | |
| **10** | Reporting blood fraction (serum, plasma, culture supernatant or whole blood) (1= Yes, 0 = No) | |
| | | **Total quality score (10 points)** |
| **Biomarker confounders red points** | | |

| *The red points should not be given if the item is statistically controlled for* | |
|---|---|
| **1** | 3 red points for comorbid illnesses such as autoimmune disorders & other immune disorders including rheumatoid arthritis, psoriasis, inflammatory bowel disease, chronic obstructive pulmonary disease, multiple sclerosis |
| **2** | 3 red points for use of recreational drugs such as methamphetamine or opioids |
| **3** | 2 red points when groups were not matched for age |
| **4** | 2 red points when groups were not matched for sex |
| **5** | 2 red points for medication use as for example immunomodulators |
| **6** | 2 red points for early traumatic life events |
| **7** | 2 red points for shift work and primary sleep disorders |
| **8** | 1.5 red points for use of antipsychotics |
| **9** | 1 red point for more common systemic immune disorders including diabetes type 1/2, essential hypertension, metabolic syndrome |
| **10** | 1 red point for not fasting (8 hours before blood extraction) |
| **11** | 1 red point for use of omega-3 and antioxidant supplements |
| **12** | 1 red point when data were not controlled for body mass index |
| **13** | 1 red point when data were not controlled for physical activity or sedentary life |
| **14** | 1 red point when data were not controlled for smoking |

| | |
|---|---|
| **15** | 1 red point for use of oral contraceptives or NSAIDs |
| **16** | 0.5 red points when data were not controlled for ethnicity in countries such as US, Brazil |
| **17** | 0.5 red points when data were not controlled for seasonality |
| **18** | 0.5 red points when data were not controlled for diurnal variation (8-10 a.m. versus all other time points) |
| | **Total red point score (26 points)** |

**ESF, Table 3.** PRISMA checklist

| Section/topic | # | Checklist item | Reported on page # |
|---|---|---|---|
| **TITLE** | | | |
| Title | 1 | Identify the report as a systematic review, meta-analysis, or both. | 1 |
| **ABSTRACT** | | | |
| Structured summary | 2 | Provide a structured summary including, as applicable: background; objectives; data sources; study eligibility criteria, participants, and interventions; study appraisal and synthesis methods; results; limitations; conclusions and implications of key findings; systematic review registration number. | 3 |
| **INTRODUCTION** | | | |
| Rationale | 3 | Describe the rationale for the review in the context of what is already known. | 5 |
| Objectives | 4 | Provide an explicit statement of questions being addressed with reference to participants, interventions, comparisons, outcomes, and study design (PICOS). | 8 |
| **METHODS** | | | |
| Protocol and registration | 5 | Indicate if a review protocol exists, if and where it can be accessed (e.g., Web address), and, if available, provide registration information including registration number. | 9 |
| Eligibility criteria | 6 | Specify study characteristics (e.g., PICOS, length of follow-up) and report characteristics (e.g., years considered, language, publication status) used as criteria for eligibility, giving rationale. | 10 |
| Information sources | 7 | Describe all information sources (e.g., databases with dates of coverage, contact with study authors to identify additional studies) in the search and date last searched. | 9 |
| Search | 8 | Present full electronic search strategy for at least one database, including any limits used, such that it could be repeated. | ESF, Table 1, page 3 |
| Study selection | 9 | State the process for selecting studies (i.e., screening, eligibility, included in systematic review, and, if applicable, included in the meta-analysis). | 10 |
| Data collection process | 10 | Describe method of data extraction from reports (e.g., piloted forms, independently, in duplicate) and any processes for obtaining and confirming data from investigators. | 11 |

| | | | |
|---|---|---|---|
| Data items | 11 | List and define all variables for which data were sought (e.g., PICOS, funding sources) and any assumptions and simplifications made. | 11 |
| Risk of bias in individual studies | 12 | Describe methods used for assessing risk of bias of individual studies (including specification of whether this was done at the study or outcome level), and how this information is to be used in any data synthesis. | 12 |
| Summary measures | 13 | State the principal summary measures (e.g., risk ratio, difference in means). | 12 |
| Synthesis of results | 14 | Describe the methods of handling data and combining results of studies, if done, including measures of consistency (e.g., $I^2$) for each meta-analysis. | 12 |
| Risk of bias across studies | 15 | Specify any assessment of risk of bias that may affect the cumulative evidence (e.g., publication bias, selective reporting within studies). | Table 3, page 71 |
| Additional analyses | 16 | Describe methods of additional analyses (e.g., sensitivity or subgroup analyses, meta-regression), if done, indicating which were pre-specified. | 12 |
| **RESULTS** | | | |
| Study selection | 17 | Give numbers of studies screened, assessed for eligibility, and included in the review, with reasons for exclusions at each stage, ideally with a flow diagram. | 14 |
| Study characteristics | 18 | For each study, present characteristics for which data were extracted (e.g., study size, PICOS, follow-up period) and provide the citations. | ESF, Table 2 page |
| Risk of bias within studies | 19 | Present data on risk of bias of each study and, if available, any outcome level assessment (see item 12). | Table 3, page 71 |
| Results of individual studies | 20 | For all outcomes considered (benefits or harms), present, for each study: (a) simple summary data for each intervention group (b) effect estimates and confidence intervals, ideally with a forest plot. | Table 1, page 66 |
| Synthesis of results | 21 | Present results of each meta-analysis done, including confidence intervals and measures of consistency. | Table 2, page 68 |
| Risk of bias across studies | 22 | Present results of any assessment of risk of bias across studies (see Item 15). | Table 3 page 71 |
| Additional analysis | 23 | Give results of additional analyses, if done (e.g., sensitivity or subgroup analyses, meta-regression [see Item 16]). | Table 2, page 68 |

| DISCUSSION | | | |
|---|---|---|---|
| Summary of evidence | 24 | Summarize the main findings including the strength of evidence for each main outcome; consider their relevance to key groups (e.g., healthcare providers, users, and policy makers). | Page 22-31 |
| Limitations | 25 | Discuss limitations at study and outcome level (e.g., risk of bias), and at review-level (e.g., incomplete retrieval of identified research, reporting bias). | Page 31-32 |
| Conclusions | 26 | Provide a general interpretation of the results in the context of other evidence, and implications for future research. | Page 32 |
| **FUNDING** | | | |
| Funding | 27 | Describe sources of funding for the systematic review and other support (e.g., supply of data); role of funders for the systematic review. | Page 33 |

**ESF, Table 4.** Studies excluded from the meta-analysis.

| Authors, year | Reason why excluded from the meta-analysis |
|---|---|
| (Meier, Drevets et al. 2018) | No data about healthy control group |
| (Møller, Kirk et al. 1980) | No baseline level of tryptophan |
| (Nazzari, Molteni et al. 2020) | No data about healthy control group |

**ESF, table 5.** Characteristics of the studies included in the systematic reviews and meta-analysis

| NO | Authors, years | Setting | Type of case | Type of Control | Sample Size | | | Age | | Assessed Biomarkers | Specimen | Method | Quality score | Red point score | Findnigs |
|---|---|---|---|---|---|---|---|---|---|---|---|---|---|---|---|
| | | | | | Cases M/F | Control M/F | Total M/F | Case-Mean (SD) | Control-Mean(SD) | | | | | | |
| 1 | Aarsland, Leskauskaite et al. 2019 | Norway | MDD | Healthy Control | 27 12/15 | 14 6/8 | 41 18/23 | 45,3(10,7) | - | TRP,KYN, KA,3HK, XA, AA, 3HA,QA,PA | Serum | LCMS | 5,75 | 13,5 | TRP*,KYN$^{\#}$, KA*,3HK*, XA*, AA*, 3HA*,QA$^{\#}$,PA* |
| 2 | Achtyes, Keaton et al. 2020 | USA | Post partum depression | Healthy Control | 87 0/87 | 60 0/60 | 147 0/147 | 26.7(5.2) | 28(6.3) | TRP, KYN, KA,QA | Plasma | HPLC-UV | 5,75 | 14 | TRP*, KYN*, KA*,QA* |
| 3 | van den Ameele, van Nuijs et al. 2020 | Belgium | BD-Depression | Healthy Control | 35 11/24 | 35 16/29 | 70 27/53 | 43.7(9.7) | 42.7(11.6) | TRP, KYN, 3HK,QA, KA | Plasma | UPLC-MS/MS | 7,75 | 10 | TRP*, KYN*, 3HK*,QA*, KA* |
| | | | BD-hypomania | Healthy Control | 32 15/17 | 35 16/29 | 67 31/46 | 42.4(12.7) | 42.7(11.6) | TRP, KYN, 3HK,QA, KA | Plasma | UPLC-MS/MS | | | TRP*, KYN*, 3HK*,QA*, KA* |
| 4 | Anderson, Parry-Billings et al. 1990 | UK | MDD | Healthy Control | 32 15/16 | 31 15/16 | 62 30/32 | 34.4(8.5) | 33.9(7.7) | TRP | Plasma | Fluorometry | 4 | 12,5 | TRP* |
| 5 | Baranyi, Amouzadeh-Ghadikolai et al. 2017 | Austria | MDD | Healthy Control | 61 20/41 | 45 14/31 | 106 34/72 | 49,8(9,6) | 44,8(17,9) | QA | Serum | LCMS | 4 | 15,5 | QA* |
| 6 | Bay-Richter, Linderholm et al. 2015 | Sweden | Affective disorders | Healthy Control | 30 9/21 | 36 29/7 | 66 38/28 | 40(8,7) | 29,7(7,3) | QA,KA | CSF | GCMS,HPLC | 3 | 18,5 | QA$^{\#}$,KA* |
| 7 | Birner, Platzer et al. 2017 | Austria | BD | Healthy Control | 143 80/63 | 101 40/61 | 244 120/124 | 43.9(13.3) | 40.3(16.4) | KYN,KA, 3HK, AA | Plasma | LC-MS/MS | 6 | 18,5 | KYN$^{\#}$,KA*, 3HK$^{\#}$, AA$^{\#}$ |
| 8 | Bradley, Case et al. 2015 | USA | MDD-Non suicidal | Healthy Control | 30 18/12 | 22 9/13 | 52 27/25 | 15.2(1.9) | 15.9(2.6) | TRP. KYN, 3HA | Plasma | HPLC | 4,25 | 16 | TRP$^{\#}$. KYN*, 3HA$^{\#}$ |
| | | | MDD-suicidal | Healthy Control | 20 5/15 | 22 9/13 | 42 14/28 | 16.8(1.8) | 15.9(2.6) | TRP. KYN, 3HA | Plasma | HPLC | | | TRP*. KYN*, 3HA* |
| 9 | Brundin, Sellgren et al. 2016 | Sweden | MDD | Healthy Control | 22 - | 36 - | 58 - | 42.1(13.6) | 32.5(11.3) | PA | CSF | GCMS | 5,5 | 18 | PA* |
| | | | Dysthymia | Healthy Control | 4 - | 36 - | 40 - | 44.3(23.3) | 32.5(11.3) | PA | CSF | GCMS | | | PA* |
| | | | Depression NOS | Healthy Control | 9 - | 36 - | 45 - | 30.8(6.6) | 32.5(11.3) | PA | CSF | GCMS | | | PA* |
| | | | MDD | Healthy Control | 15 - | 29 - | 44 - | 43.1(13.5) | 40.6(11.5) | PA, QA | Plasma | GCMS | | | PA*, QA$^{\#}$ |
| | | | Dysthymia | Healthy Control | 3 - | 29 - | 32 - | 43.0(18.9) | 40.6(11.5) | PA, QA | Plasma | GCMS | | | PA*, QA* |

| | | | | | | | | | | | | | | | |
|---|---|---|---|---|---|---|---|---|---|---|---|---|---|---|---|
| | | | BD | Healthy Control | 21<br>- | 29<br>- | 50<br>- | 39.2(12.4) | 40.6(11.5) | PA, QA | Plasma | GCMS | | | PA*, QA$^{\#}$ |
| | | | Depression UNS | Healthy Control | 4<br>- | 29<br>- | 33<br>- | 28.1(8.5) | 40.6(11.5) | PA, QA | Plasma | GCMS | | | PA*, QA# |
| 10 | Busse, Busse et al. 2015 | Germany | Depression | Healthy Control | 6<br>2/4 | 10<br>5/5 | 16<br>7/9 | 47(10) | 56(6) | QA | Brain Tissue | - | 2,75 | 16 | QA* |
| 11 | Carrillo-Mora, Pérez-De la Cruz et al. 2020 | Mexico | Poststroke depression | Healthy Control | 38<br>21/17 | 22<br>14/8 | 60<br>35/25 | 57.3(13,4) | 57,9(14,7) | 3HK, KA | Serum | HPLC | 6,25 | 10,25 | 3HK$^{\#}$, KA$^{\#}$ |
| 12 | Castillo, Murata et al. 2019 | USA | Bipolar-Depression | Healthy Control | 55<br>27/28 | 38<br>16/22 | 93<br>43/50 | 52.9(9.8) | 40.6(10.2) | QA | Serum | UPLC-MS | 6 | 13,5 | QA* |
| 13 | Cathomas, Guetter et al. 2021 | Switzerland | MDD | Healthy Control | 43<br>17/26 | 19<br>9/10 | 62<br>26/36 | 35,7(10,8) | 32,5(9,4) | KYN,KA,3HK,QA,TRP | Plasma | HPLC | 5 | 16 | KYN*,KA*,3HK*, QA*,TRP* |
| 14 | Chiaroni, Azorin et al. 1990 | Switzerland | DD | Healthy Control | 21<br>11/10 | 33<br>14/19 | 54<br>25/29 | 41.5(11.5) | 39.4(8.3) | TRP | Plasma | HPLC | 5 | 16,5 | TRP* |
| | | | MRD | | 25<br>6/19 | | 58<br>20/38 | 59.04(11.4) | | | | | | | TRP* |
| | | | BD | | 18<br>7/11 | | 51<br>21/30 | 58.1(12.3) | | | | | | | TRP* |
| 15 | Cho, Savitz et al. 2017 | USA | MDD | Healthy Control | 68<br>13/58 | 66<br>24/42 | 134<br>37/97 | 36,2(9,9) | 32,3(10,2) | TRP,KYN, KA,3HK,QA | Serum | HPLC MS/MS | 5,25 | 15 | TRP*,KYN*, KA*,3HK$^{\#}$,QA* |
| | | | Previous MDD | | 26<br>9/17 | | 92<br>33/59 | 33,3(11,1) | | | | | | | TRP*,KYN*, KA*,3HK*,QA$^{\#}$ |
| 16 | Clark, Pocivavsek et al. 2016 | USA | Depression | Healthy Control | 45<br>30/15 | 36<br>27/9 | 91<br>57/24 | 43.3(13.6) | 42.1(14.3) | TRP,KYN, KA,3-HK, QA | Brain Tissue | HPLC,GC/MS | 2,75 | 17,5 | TRP*,KYN*, KA*,3-HK*, QA* |
| 17 | Colle, Masson et al. 2020 | France | MDE | Healthy Control | 173<br>61/112 | 214<br>90/124 | 387<br>150/237 | 48 | 45,5 | TRP,AA,KA KYN,PA,XA 3HA | Plasma | UPLC-MS | 5 | 11,5 | TRP$^{\#}$,AA*,KA* KYN*,PA*,XA* 3HA* |
| 18 | Coppen, Brooksbank et al. 1972 | UK | MDD | Healthy Control | 10<br>2/8 | 14<br>7/7 | 24<br>9/15 | 49,7(16,4) | 50,4(13,3) | TRP | CSF | Fluorometry | 1 | 21 | TRP* |
| | | | Mania | | 3<br>1/2 | | 17<br>8/9 | 43,6(17,6) | | | | | | | TRP* |
| 19 | Coppen, Eccleston et al. 1973 | UK | UD | Healthy Control | 24<br>0/24 | 26<br>0/26 | 50<br>0/50 | 55,3(13,7) | 48,6(11,2) | TRP | Plasma | Fluorometry | 4,75 | 15 | TRP* |
| 20 | Cowen, Parry-Billings et al. 1989 | UK | MDD | Healthy Control | 12<br>6/6 | 12<br>6/6 | 24<br>12/12 | 36,5<br>(26-55) | 36,2<br>(28-54) | TRP | Plasma | Fluorometry | 6,75 | 11 | TRP* |
| 21 | Czermak, Hauger et al. 2008 | USA | MDD-remitted | Healthy Control | 28<br>8/20 | 26<br>8/18 | 54<br>16/38 | 39,5(12,5) | 33,8(11,2) | TRP | Plasma | HPLC | 4,5 | 12,5 | TRP* |
| 22 | Dahl, Andreassen et al. 2015 | Norway | MDD | Healthy Control | 50<br>12/38 | 34<br>15/19 | 84<br>27/57 | 40(12) | 38,3(13,9) | TRP,KYN, KA, QA | Plasma | HPLC | 4,25 | 15 | TRP$^{\#}$,KYN*, KA$^{\#}$, QA* |
| 23 | DeMyer, Shea et al. 1981 | USA | Depression | Healthy Control | 18<br>5/13 | 10<br>3/7 | 28<br>8/20 | 33.4 | 33.3 | TRP | Plasma | Fluorometry | 4,5 | 14 | TRP* |

| | | | | | | | | | | | | | | | |
|---|---|---|---|---|---|---|---|---|---|---|---|---|---|---|---|
| 24 | DeWitt, Bradley et al. 2018 | USA | MDD | Healthy Control | 14<br>3/11 | 7<br>3/4 | 21<br>6/15 | 16,7(3,04) | 16,53(1,7) | KYN/TRP, KA, QA | Plasma | LCMS/MS | 3,75 | 16,5 | KYN/TRP#, KA*, QA* |
| 25 | Doolin, Allers et al. 2018 | Ireland | First presentation MDD | Healthy Control | 39<br>16/23 | 37<br>18/19 | 76<br>43/42 | 24.3(7.6) | 30.8(10.7) | TRP, KYN QA, KA | Plasma | LC–MS/MS | 5 | 9 | TRP*, KYN* QA*, KA* |
| | | | Remitted-MDD | | 35<br>11/24 | 37<br>18/19 | 72<br>29/43 | 42.4(11.5) | | | | | | | TRP*, KYN* QA#, KA* |
| 26 | Ebesunun, Eruvulobi et al. 2012 | Nigeria | MDD | Healthy Control | 30<br>9/21 | 30<br>- | 60 | 37,9(2) | 42,5(1) | TRP | Plasma | HPLC | 4,25 | 15 | TRP* |
| 27 | Erabi, Okada et al. 2020 | Japan | MDD | Healthy Control | 88<br>46/42 | 88<br>40/48 | 176<br>86/90 | 42,9(12,2) | 41,3(12,7) | TRP, KYN, 3HK | Plasma | LCMS | 5,25 | 15 | TRP*, KYN*, 3HK* |
| 28 | Erhardt, Lim et al. 2013 | Sweden | MDD | Healthy Control | 29 | 7 | 36 | - | 30(18-66) | KA, QA | CSF | HPLC-GCMS | 4,5 | 15 | KA*, QA# |
| 29 | Gabbay, Klein et al. 2010 | USA | MDD-M | Healthy Control | 20<br>9/11 | 22<br>9/13 | 42<br>18/24 | 15.6(2.0) | 16.0(2.7) | TRP, KYN | Plasma | HPLC | 6,25 | 9,5 | TRP*, KYN* |
| | | | MDD-non M | | 30<br>14/16 | | 52<br>23/29 | 16.3(2.1) | | | | | | | TRP#, KYN* |
| 30 | Georgin-Lavialle, Moura et al. 2016 | France | Depressive symptoms | Healthy Control | 54 | 54 | 108 | 50,1 | 50 | TRP, KYN,QA, KA | Plasma | HPLC | 3 | 18 | TRP*, KYN*,QA#, KA# |
| 31 | Gerner, Fairbanks et al. 1984 | USA | Depression | Healthy Control | 41<br>18/23 | 38<br>18/20 | 79<br>36/43 | 40.3(11.07) | 33.4(11.7) | TRP | CSF | LC | 3,5 | 16 | TRP* |
| | | | Mania | | 15<br>5/10 | | 51<br>28/5 | 35.7(11.4) | | | | | | | TRP* |
| 32 | Guicheney, Léger et al. 1988 | France | Depressive symptoms | Healthy Control | 31 | 14 | 45 | - | - | TRP | Plasma | | 5 | 13,5 | TRP* |
| 33 | Manjarrez-Gutierrez, Marquez et al. 2009 | Mexico | MDD | Healthy Control | 8<br>4/4 | 9<br>4/5 | 17<br>8/9 | 14.8(0.6) | 14.2(2.1) | TRP | Plasma | HPLC | 6 | 10,5 | TRP* |
| | | | MDD-DM | | 9<br>6/3 | | 18<br>10/8 | 14.4(1.6) | | | | | | | TRP* |
| 34 | Hayward, Goodwin et al. 2005 | UK | Euthymia | Healthy Control | 24<br>10/14 | 24<br>10/14 | 48<br>20/28 | 38.8(3.5) | 37.4(3.9) | TRP | Plasma | HPLC | 6 | 12,5 | TRP# |
| 35 | Healy, Carney et al. 1982 | Ireland | Depression | Healthy Control | 18<br>- | 20<br>- | 38<br>- | 40.02(12.1) | 32 | TRP | Plasma | Ultrafiltration apparatus | 4 | 14,5 | TRP# |
| 36 | Heilman, Hill et al. 2019 | USA | Affective disorder | Healthy Control | 96<br>41/55 | 56<br>27/29 | 152<br>68/84 | 35,8(7,5) | 31,3(8,6) | TRP,KYN, KA,QA,PA | Plasma | HPLC | 6,25 | 12 | TRP*,KYN*, KA#,QA*,PA# |
| 37 | Hennings, Schwarz et al. 2013 | Germany | MDD | Healthy Control | 38<br>15/23 | 48<br>16/32 | 86<br>31/55 | 32,08(12,2) | 36,4(13,2) | TRP, KYN | Serum | HPLC | 5 | 13 | TRP#, KYN* |
| 38 | Hoekstra, van den Broek et al. 2001 | Netherland | MDD | Healthy Control | 20<br>7/13 | 29<br>13/16 | 49<br>23/26 | 52(13,1) | 37(8,3) | TRP | Plasma | HPLC | 3 | 19 | TRP* |

| | | | | | | | | | | | | | | | |
|---|---|---|---|---|---|---|---|---|---|---|---|---|---|---|---|
| 39 | Hoekstra, Fekkes et al. 2006 | Netherland | BD | Healthy Control | 20<br>14/6 | 20<br>14/6 | 40<br>28/12 | 50,2(13,8) | 50,1(13,5) | TRP | Plasma | HPLC | 3,75 | 15 | TRP* |
| 40 | Hu, Li et al. 2017 | China | MDD | Healthy Control | 54 | 26 | 80 | - | - | TRP,KYN, KA | Serum | HPLC MS/MS | 4,25 | 20,5 | TRP*,KYN*, KA* |
| 41 | Hüfner, Oberguggenberger et al. 2015 | Austria | Depression | Healthy Control | 29 | 28 | 57 | 53,9(1,6) | 42,9(2,1) | KYN/TRP | Serum | HPLC | 6 | 13 | KYN/TRP* |
| | | | | | 46 | | 74 | 43,9(1,6) | | | | | | | KYN/TRP# |
| 42 | Hughes, Carballedo et al. 2012 | Ireland | MDD | Healthy Control | 39<br>16/23 | 39<br>17/22 | 78<br>33/45 | 41,9(11,2) | 37,1(13,1) | TRP,KYN, KA,3HA | Plasma | HPLC | 5,5 | 12,5 | TRP*,KYN*, KA*,3HA* |
| 43 | Joseph, Brewerton et al. 1984 | USA | MDD | Healthy Control | 8<br>3/5 | 8<br>3/5 | 16<br>6/10 | 45(17) | 42(16) | TRP | Plasma | HPLC | 6,5 | 9 | TRP* |
| | | | | | 8<br>3/5 | 8<br>3/5 | 16<br>6/10 | 42(13) | | | | | | | TRP* |
| 44 | Karege, Widmer et al. 1994 | Switzerland | MDD | Healthy Control | 30<br>10/20 | 20<br>10/10 | 50<br>20/30 | 47(10) | 47(9) | TRP | Plasma | HPLC | 6 | 12,5 | TRP* |
| 45 | Krause, Myint et al. 2017 | Germany | MDD | Healthy Control | 32<br>16/16 | 20<br>15/5 | 52<br>31/21 | 44,6(11,6) | 40(10,4) | KYN,KA | Serum | HPLC | 6,75 | 9 | KYN*,KA* |
| 46 | Krause, Kirnich et al. 2019 | Germany | MDD | Healthy Control | 39<br>15/24 | 51<br>18/33 | 90<br>33/57 | 32,6(12,5) | 36,5(13,1) | TRP, KYN, KA, 3HK | Serum | HPLC | 7 | 4,5 | TRP*, KYN*, KA#, 3HK* |
| 47 | Kuwano, Kato et al. 2018 | Japan | MDD | Healthy Control | 15<br>9/6 | 19<br>10/9 | 34<br>19/15 | 30.1(7.6) | 30.2(6.8) | TRP,KYN, KA,XA | Plasma | LC-MS | 4,75 | 12,5 | TRP*,KYN*, KA*,XA* |
| 48 | Liu, Ding et al. 2018 | China | MDD | Healthy Control | 33<br>10/23 | 23<br>12/11 | 56<br>22/34 | 28,3(5,2) | 29,3(5,8) | TRP,KYN, KA, QA | Plasma | UPLC-3Q-MS | 6,75 | 9,5 | TRP*,KYN*, KA*, QA* |
| | | | BD | | 20<br>8/12 | | 43<br>20/23 | 30,4(5,08) | | | | | | | TRP*,KYN*, KA*, QA* |
| 49 | Lucca, Lucini et al. 1992 | Italy | BD | Healthy Control | 9<br>4/5 | 29<br>15/14 | 38<br>19/19 | 44.2(9.7) | 40,1(9,3) | TRP | Plasma | | 5 | 13 | TRP* |
| | | | MDD | | 19<br>7/12 | | 48<br>22/26 | 44,5(11,7) | | | | | | | TRP* |
| 50 | Maes, Jacobs et al. 1990 | Belgium | MDD | Healthy Control | 15<br>6/9 | 16<br>8/8 | 31<br>14/17 | 41.0(15.8) | 40.1(10.1) | TRP | Plasma | LC | 6 | 10,5 | TRP* |
| | | | | | 22<br>10/12 | | 38<br>18/20 | 43.9(13.8) | | | | | | | TRP* |
| | | | | | 13<br>6/7 | | 29<br>14/15 | 52.2(14.5) | | | | | | | TRP* |
| 51 | Maes, Meltzer et al. 1993 | Belgium | MDD | Healthy Control | 7 | 8 | 15 | 33.0(13.1) | 44.1(13.1) | TRP | Plasma | LC | 6,75 | 9,5 | TRP* |
| | | | | | 7 | 8 | 15 | 51.7(17.0) | | | | | | | TRP* |
| | | | | | 10 | 8 | 18 | 54.8(12.5) | | | | | | | TRP* |
| 52 | Maes, De Backer et al. 1995 | Belgium | MDD | Healthy Control | 41<br>17/24 | 50<br>26/24 | 91<br>43/48 | 42.1(14.2) | 40.7(11.3) | TRP | Plasma | LC | 6,25 | 12 | TRP* |
| | | | | | 47<br>12/35 | | 97<br>38/59 | 47.3(13.5) | | | | | | | TRP* |
| | | | | | 35<br>14/21 | | 85<br>40/45 | 51.7(15.0) | | | | | | | TRP* |
| 53 | | Belgium | MDD | | 12 | 24 | 36 | 42,8(11,9) | 41,1(13,5) | TRP | Serum | HPLC | 5,75 | 11,5 | TRP* |

| | | | | | | | | | | | | | | | |
|---|---|---|---|---|---|---|---|---|---|---|---|---|---|---|---|
| | | | | | 4/8 | 11/13 | 15/21 | | | | | | | | |
| | Maes, Wauters et al. 1996 | | | Healthy Control | 21<br>3/18 | | 45<br>14/31 | 48,7(14,3) | | | | | | | TRP* |
| | | | | | 9<br>3/6 | | 33<br>14/19 | 58,7(14) | | | | | | | TRP* |
| 54 | Maes, Galecki et al. 2011 | Germany | MDD | Healthy Control | 39<br>10/29 | 35<br>17/18 | 74<br>27/47 | 40,2(10,4) | 38(11.9) | TRP, KYN, KA | Plasma | HPLC | 8 | 7,5 | TRP*, KYN*, KA* |
| | | | | | 36<br>13/23 | | 71<br>30/41 | 41,9(9,9 | | | | | | | TRP*, KYN*, KA* |
| 55 | Maes and Rief 2012 | Germany | MDD | Healthy Control | 35<br>13/22 | 22<br>14/8 | 57<br>27/30 | 42,1(10) | 39(13) | TRP, KYN/TRP, KYN/KA | Plasma | HPLC | 8 | 7,5 | TRP*, KYN/TRP#, KYN/KA# |
| | | | | | | | | 40,2(10,4) | | | | | | | TRP*, KYN/TRP#, KYN/KA# |
| 56 | Maes, Verkerk et al. 1997 | Belgium | MDD | Healthy Control | 35<br>17/18 | 15<br>10/5 | 50<br>27/23 | 50,7(14,4) | 47,5(15) | TRP | Serum | HPLC | 6,75 | 9 | TRP* |
| 57 | Mauri, Ferrara et al. 1998 | Italy | MDD | Healthy Control | 29<br>15/14 | 28<br>16/12 | 57<br>31/26 | 47.4(10.8) | 42.4(14.1) | TRP | Plasma | LC | 5 | 11 | TRP* |
| 58 | Mauri, Boscati et al. 2001 | Italy | MDD | Healthy Control | 16<br>5/11 | 11<br>9/2 | 27<br>14/13 | 50.1(11.5) | 39.9(13.3) | TRP | Plasma | LC | 5 | 13,5 | TRP* |
| 59 | Meier, Drevets et al. 2016 | USA | MDD | Healthy Control | 73<br>16/57 | 91<br>36/55 | 164<br>52/112 | 34,2(9,3) | 31,6(9,3) | TRP,KYN, KA,3HK,QA | Serum | HPLC MS/MS | 7,25 | 10 | TRP*,KYN*, KA*,3HK#,QA# |
| 60 | Menna-Perper 1983 | USA | MDD-M | Healthy Control | 9<br>6/3 | 6<br>3/3 | - | - | - | TRP | Plasma | - | - | - | TRP* |
| 61 | Milaneschi, Allers et al. 2021 | Netherland | MDD-remitted | Healthy Control | 753<br>226/527 | 642<br>246/396 | 1395<br>472/923 | 43,5(12,5) | 41,1(14,7) | TRP,KYN, KA, QA | Plasma | - | 3 | 17,5 | TRP*,KYN*, KA*, QA* |
| | | | MDD-current | | 1100<br>358/743 | | 1742<br>603/1139 | 40,8(12,1) | | | | | | | TRP#,KYN*, KA*, QA* |
| 62 | Miller, Llenos et al. 2006 | USA | MDD | Healthy Control | 14 | 14 | 28 | 45.9(9.7) | 48.6(10.8) | KYN, KA | Brain Tissue | HPLC | 4,75 | 12,5 | KYN#, KA# |
| | | | BD | | 14 | 14 | 28 | 41.8(11.9) | | | | | | | KYN#, KA# |
| 63 | Moaddel, Shardell et al. 2018) | USA | MDD | Healthy Control | 29 | 25 | 54 | - | - | TRP, KYN | Plasma | HPLC | 5 | 12,5 | TRP*, KYN* |
| 64 | Møller 1993 | Denmark | Depression | Healthy Control | 26<br>8/18 | 55<br>16/39 | 81<br>24/57 | 52(10) | 38(13) | TRP | Plasma | IEC | 3,25 | 17 | TRP* |
| 65 | Moller, Kirk et al. 1976 | Denmark | Depression | Healthy Control | 19<br>2/17 | 23<br>2/21 | 42<br>4/38 | 54,1(21,8) | 48,8(15,7) | TRP, KYN | Plasma | IEC | 5 | 15 | TRP*, KYN* |
| 66 | Møller, Kirk et al. 1982 | Denmark | Depression | Healthy Control | - | - | - | 44(14) | 40(11) | TRP | Plasma | IEC | 7,75 | 9,5 | TRP* |
| 67 | Moreno, Gelenberg et al. 1999 | Mexico | MDD | Healthy Control | 12<br>4/8 | 12<br>4/8 | 24<br>8/16 | 48(19) | 47(18) | TRP | Plasma | HPLC-F | 5,5 | 9 | TRP* |
| 68 | Moreno, Gaspar et al. 2013 | USA | MDD | Healthy Control | 30<br>6/24 | 30<br>6/24 | 60<br>12/48 | 30,2(10,6) | 34,5(7,7) | TRP | Plasma | HPLC | 7 | 6,5 | TRP* |
| 69 | Mukherjee, Krishnamurthy et al. 2018 | USA | BD | Healthy Control | 21<br>11/10 | 28<br>12/16 | 49<br>23/26 | 36.1(11.3) | 31.5(10.3) | TRP, KYN | Plasma | HPLC | 3,75 | 13 | TRP*, KYN* |

| | | | | | | | | | | | | | | |
|---|---|---|---|---|---|---|---|---|---|---|---|---|---|---|
| 70 | Murata, Murphy et al. 2020 | USA | TRBDD | Healthy Control | 47<br>28/19 | 41<br>28/13 | 88<br>56/32 | 42.3(12.3) | 39.6(13.5) | KYN/TRP | Serum | HPLC | 5,75 | 10,5 | KYN/TRP* |
| 71 | Myint, Kim et al. 2007a | South Korea | BD | Healthy Control | 39<br>15/24 | 80<br>40/40 | 119<br>55/64 | 37.6(11.6) | 39.0(8.7) | TRP, KYN, KA, 3HA | Plasma | HPLC | 6 | 12 | TRP*, KYN*, KA*, 3HA$^{\#}$ |
| 72 | Myint, Kim et al. 2007b | South Korea | Depression | Healthy Control | 58<br>48/10 | 189<br>32/157 | 247<br>80/167 | 44.6(14.6) | 32.4(10.6) | TRP, KYN, KA, 3HA | Plasma | HPLC | 5,75 | 15 | TRP*, KYN*, KA*, 3HA$^{\#}$ |
| 73 | Nikkheslat, Zunszain et al. 2015 | UK | Depression | Healthy Control | 15 | 29 | 44 | - | - | TRP, KYN, KA,3HK, 3HA | Serum | HPLC | 4,25 | 15 | TRP*, KYN*, KA*,3HK*, 3HA* |
| 74 | Ogawa, Koga et al. 2018 | Japan | MDD | Healthy Control | 147<br>74/73 | 217<br>100/117 | 364<br>174/190 | 42,1(12,1) | 41,2(13,9) | TRP | Plasma | HPLC | 5,5 | 14,5 | TRP* |
| 75 | Olsson, Samuelsson et al. 2010 | Sweden | BD | Healthy Control | 31<br>31/0 | 23<br>23/0 | 54<br>54/0 | 36.3(9.2) | 33.1(6.9) | KA | CSF | HPLC | 4,75 | 14,5 | KA$^{\#}$ |
| 76 | Ortiz, Mariscot et al. 1993 | Spain | MDD | Healthy Control | 10 | 10 | 20 | 39,8(9,1) | - | TRP | Plasma | HPLC | 4 | 16 | TRP* |
| | | | | | 7 | | 17 | 68,2(4,8) | | | | | | | TRP* |
| 77 | Öztürk, Yalın Sapmaz et al. 2021 | Turkey | MDD | Healthy Control | 48<br>3/45 | 31<br>1/30 | 79<br>4/75 | 15.2(1.4) | 14.8(1.3) | TRP,KYN, KA, QA | Serum | ELISA | | | TRP*,KYN$^{\#}$, KA$^{\#}$, QA$^{\#}$ |
| 78 | Pan, Xia et al. 2018 | China | MDD | Healthy Control | 50<br>24/26 | 50<br>25/25 | 100<br>49/51 | 36.9(9.1) | 38.3(11.3) | TRP, KYN, 3HA | Plasma | LC-MS/MS | 4,5 | 14 | TRP*, KYN*, 3HA$^{\#}$ |
| | | | | | 49<br>23/26 | 40<br>22/18 | 89<br>45/44 | 37.7(11.9) | 36.8(10.1) | KYN | Plasma | GC-MS | | | KYN* |
| | | | BD | | 30<br>13/17 | | 70<br>35/35 | 35.8(10.7) | | | | | | | KYN* |
| 79 | Paul, Schwieler et al. 2022 | Sweden | MDD | Healthy Control | 63<br>19/44 | 48<br>14/34 | 111<br>33/78 | 27.5(17.2) | 32(24) | KYN, KA,3HK,QA ,PA | Plasma | UPLC-MS/MS | 6 | 12 | KYN$^{\#}$, KA*,3HK*,QA$^{\#}$,PA * |
| | | | | | 36<br>11/25 | 33<br>11/22 | 69<br>22/47 | 31(18.3) | 26(17) | | CSF | | | | KYN*, KA$^{\#}$,3HK$^{\#}$,QA#,PA * |
| 80 | Pinto, de Souza et al. 2012 | Brazil | BD | Healthy Control | 5 | 5 | 10 | 34(17.4) | 34(13.07) | TRP | Plasma | HPLC | 4,75 | 9 | TRP* |
| 81 | Platzer, Dalkner et al. 2017 | Austria | BD | Healthy Control | 42<br>42/0 | 36<br>36/0 | 78<br>78/0 | 43.6(14.3) | 38.1(15.1) | TRP,KYN, KA, 3HK | Serum | UPLC-MS/MS | 3 | 19,5 | TRP*,KYN$^{\#}$, KA*, 3HK$^{\#}$ |
| | | | | | 26<br>0/26 | 57<br>0/57 | 83<br>0/83 | 47.0(13.5) | 39.4(16.9) | | | | | | TRP*,KYN*, KA*, 3HK$^{\#}$ |
| 82 | Poletti, Melloni et al. 2019 | Italy | BD | Healthy Control | 55<br>20/35 | 36<br>13/23 | 91<br>33/58 | 47.2(9.8) | 43.8(12.2) | TRP, KYN | Plasma | HPLC | 6 | 10,5 | TRP*, KYN$^{\#}$ |
| | | | Mania | | 17<br>9/8 | | 53<br>22/31 | 50.3(11.9) | 43.8(12.2) | | | | | | TRP*, KYN$^{\#}$ |
| 83 | Poletti, Myint et al. 2018 | Italy | BD | Healthy Control | 22<br>8/14 | 14<br>6/8 | 36<br>14/22 | 46.5(13.6) | 27.2(8.3) | TRP, KYN 3HK, KA | Plasma | HPLC | 4,75 | 12,5 | TRP*, KYN* 3HK$^{\#}$, KA* |
| 84 | Pompili, Lionetto et al. 2019 | Italy | MDD | Healthy Control | 49<br>24/25 | 78<br>34/44 | 127<br>58/69 | 36,6(8,5) | 40,4(12,3) | TRP, KYN AA, XA | Plasma | LCMS | 3 | 18,5 | TRP*, KYN AA, XA |

| | | | | | | | | | | | | | | | |
|---|---|---|---|---|---|---|---|---|---|---|---|---|---|---|---|
| 85 | Porter, Gallagher et al. 2003 | UK | MDD | Healthy Control | 20<br>9/11 | 20<br>10/10 | 40<br>19/21 | 34(11) | 33(12) | TRP | Plasma | HPLC | 5,75 | 13,5 | TRP* |
| 86 | Price, Charney et al. 1991 | USA | Depression | Healthy Control | 126<br>38/88 | 58<br>17/41 | 184<br>58/126 | 43(14) | 38(13) | TRP | Serum | HPLC | 7 | 15 | TRP* |
| 87 | Quak, Doornbos et al. 2014 | Netherland | MDD | Healthy Control | 1042 | 1007 | 2049 | 40,9(12,1) | - | KYN/TRP | Plasma | XLC-MS/MS | 6 | 9 | KYN/TRP* |
| 88 | Quintana 1992 | Spain | MDD | Healthy Control | 25<br>10/15 | 25<br>10/15 | 50<br>20/30 | 54(9,3) | - | TRP | Plasma | - | 5,75 | 13,5 | TRP* |
| 89 | Reininghaus, McIntyre et al. 2014 | Austria | BD | Healthy Control | 78 | 156 | 234 | 46.8(14.07 | 35.2(11.5) | KYN, KYN/TRP | Serum | HPLC | 5,25 | 16,5 | KYN#, KYN/TRP# |
| 90 | Rief, Pilger et al. 2004 | Germany | MDD | Healthy Control | 40<br>10/30 | 36<br>18/18 | 76<br>28/48 | 40,3(10,3) | 37,8(11,8) | TRP | Plasma | - | 6,75 | 13,5 | TRP* |
| | | | | | 36<br>13/23 | | 72<br>31/41 | 42,3(10,3) | | | | | | | TRP* |
| 91 | Roiser, Levy et al. 2009 | UK | MDD | Healthy Control | 23<br>6/17 | 20<br>7/13 | 43<br>13/30 | 32,4(9,8) | 30,5(7,3) | TRP | Plasma | - | 3,75 | 11,5 | TRP# |
| 92 | Russ, Ackerman et al. 1990 | USA | Depression | Healthy Control | 16<br>6/10 | 9<br>2/7 | 25<br>8/17 | 34(13) | 37(9) | TRP | Plasma | Fluorome try | 4,75 | 23 | TRP* |
| 93 | Ryan, Allers et al. 2020 | Ireland | Depression | Healthy Control | 94<br>36/58 | 57<br>20/37 | 151<br>56/95 | 55.4(14.7) | 51.7(11.4) | TRP, KYN KA, AA, 3HK, XA 3HA, QA PA | Plasma | LC/MS | 6 | 13,5 | TRP*, KYN* KA*, AA*, 3HK*, XA* 3HA*, QA* PA* |
| 94 | Sakurai, Yamamoto et al. 2020 | Japan | Depression | Healthy Control | 42<br>42/0 | 38<br>38/0 | 80<br>80/0 | 46(6,6) | 46,1(7,4) | KYN,TRP 3HA,KA AA,3HK | Serum | HPLC | 5 | 15,5 | KYN*,TRP* 3HA#,KA# AA#,3HK* |
| | | | | | 19<br>0/19 | 21<br>0/21 | 40<br>0/40 | 46,5(8,1) | 43,3(7,3) | | | | | | KYN*,TRP* 3HA*,KA# AA#,3HK# |
| 95 | Savitz, Dantzer et al. 2015a | USA | BD | Healthy Control | 38<br>8/30 | 48<br>19/29 | 86<br>27/59 | 40.5(11.7) | 32.6(10.3) | TRP, KYN KA, 3HK QA | Serum | HPLC MS/MS | 5 | 14,5 | TRP*, KYN* KA*, 3HK* QA# |
| | | | | | 25<br>4/21 | | 73<br>23/50 | 36.2(10.5) | | | | | | | TRP*, KYN* KA*, 3HK* QA# |
| 96 | Savitz, Dantzer et al. 2015b | USA | MDD | Healthy Control | 29<br>5/24 | 20<br>10/10 | 49<br>15/34 | 36,4(10) | 35(10,9) | KYN, KA 3HK, QA TRP | Serum | HPLC | 6,75 | 9,5 | KYN#, KA* 3HK#, QA# TRP* |
| 97 | Savitz, Dantzer et al. 2015c | USA | MDD-remitted | Healthy Control | 21<br>9/12 | 58<br>25/33 | 79<br>34/45 | 30,8(12,2) | 32,8(10,7) | TRP, KYN KA, 3HK QA | Serum | HPLC MS/MS | 6,75 | 9,5 | TRP*, KYN# KA*, 3HK* QA* |
| | | | MDD-current | Healthy Control | 49<br>11/38 | | 107 | 35,4(9,8) | 32,8(10,)7) | | | | | | TRP*, KYN# KA*, 3HK# QA# |
| 98 | Savitz, Drevets et al. 2015d | USA | MDD | Healthy Control | 53<br>11/42 | 47<br>18/29 | 100<br>29/71 | 34.6(9.8) | 34.3(11.4) | TRP, KYN | Serum | LC–MS/MS | 7 | 11,5 | TRP*, KYN* KA*, 3HK#, QA# |

| | | | | | | | | | | | | | | | |
|---|---|---|---|---|---|---|---|---|---|---|---|---|---|---|---|
| | | | | | | | | | | KA, 3HK,<br>QA | | | | | |
| 99 | Schwieler, Samuelsson et al. 2016 | Sweden | MDD | Healthy Control | 19<br>11/8 | 14<br>14/0 | 33<br>25/8 | 39.9(23.2) | 45.6(20.3) | KA, TRP<br>KYN, QA | Plasma | HPLC | 6,25 | 19 | KA*, TRP*<br>KYN*, QA$^{\#}$ |
| 100 | Sellgren, Gracias et al. 2019 | Sweden | BD | Healthy Control | 163<br>64/99 | 114<br>52/62 | 277<br>116/161 | 34.2(3.3) | 35.1(3.1) | KA | Plasma | HPLC | 6 | 13,5 | KA* |
| | | | | | 94<br>39/55 | 113<br>51/62 | 207<br>90/117 | 36(20.5) | - | | CSF | | | | KA$^{\#}$ |
| 101 | Shaw, Tidmarsh et al. 1980 | UK | UD | Healthy Control | 75<br>27/48 | 36<br>17/19 | 111<br>44/67 | - | - | TRP | Plasma | kineticall<br>y | 2 | 19 | TRP* |
| | | | BD | | 22<br>10/12 | | 58<br>27/31 | | | | | | | | TRP* |
| 102 | Song, Lin et al. 1998 | Belgium | Depression | Healthy Control | 6<br>2/4 | 14<br>6/8 | 20<br>8/12 | 50.3(15.3) | 45.5(15.5) | TRP | Plasma | HPLC | 4,25 | 13,5 | TRP* |
| 103 | Sorgdrager, Doornbos et al. 2017 | Netherland | MDD | Healthy Control | 625<br>202/423 | 406<br>163/243 | 1031<br>365/666 | 42.7(12.8) | 42.9(14.7) | TRP, KYN | Serum | XLC-MS/MS | 5,5 | 15,5 | TRP$^{\#}$, KYN* |
| | | | | | 695<br>215/480 | | 1101<br>378/723 | 44.3(11.7) | | | | | | | TRP$^{\#}$, KYN* |
| 104 | Steen, Dieset et al. 2020 | USA | BD | Healthy Control | 73<br>33/40 | 68<br>33/35 | 141<br>66/75 | 31.0(19.0) | 31.0(16.0) | 3HK, XA | Plasma | LCECA | 4,25 | 14,5 | 3HK*, XA* |
| 105 | Steiner, Walter et al. 2011 | Germany | Depression | Healthy Control | 12<br>6/6 | 10<br>5/5 | 22<br>11/11 | 51(9) | 56(6) | QA | Brain Tissue | ELISA | 3,75 | 16 | QA$^{\#}$ |
| 106 | Sublette, Galfalvy et al. 2011 | USA | MDD | Healthy Control | 30<br>16/14 | 31<br>10/21 | 61<br>26/35 | 37.8(13.08 | 35.6(13.9) | TRP, KYN | Serum | HPLC | 4 | 14 | TRP*, KYN$^{\#}$ |
| 107 | Sun, Drevets et al. 2020 | Canada | MDD | Healthy Control | 161<br>62/99 | 28<br>9/19 | 189<br>71/118 | 40.01(12.0<br>4) | 46.5(14.5) | TRP, KYN<br>3HK, KA<br>QA | Plasma | LC-MS | 4,75 | 18 | TRP*, KYN<br>3HK*, KA<br>QA* |
| 108 | Teraishi, Hori et al. 2015 | Japan | MDD | Healthy Control | 18<br>8/10 | 24<br>12/12 | 42<br>20/22 | 39,9(10,1) | 45,1(11,9) | TRP | Plasma | - | 4 | 13 | TRP* |
| 109 | Trepci, Sellgren et al. 2021 | Sweden | BD | Healthy Control | 101<br>40/61 | 80<br>39/41 | 181<br>79/102 | 44.2(13.9) | 34.7(12.4) | TRP, KYN<br>KA, PA, QA | CSF | UPLC-MS/MS | 7 | 16,5 | TRP*, KYN$^{\#}$<br>KA$^{\#}$, PA$^{\#}$, QA$^{\#}$ |
| 110 | Umehara, Numata et al. 2017 | Japan | MDD | Healthy Control | 33<br>10/23 | 33<br>10/23 | 66<br>20/46 | 47,1(13,2) | 46,5(10,1) | KYN,<br>KYN/TRP | Plasma | CE-TOFMS | 6,75 | 12,5 | KYN*,<br>KYN/TRP* |
| 111 | Veen, Myint et al. 2016 | Netherland | Post Partum depression | Healthy Control | 29<br>0/29 | 29<br>0/29 | 58<br>0/58 | 32.9(4.6) | 25.04(5.4) | TRP,KYN,<br>3HK,QA,KA | Serum | LC–MS/MS | 5 | 16 | TRP#,KYN*,<br>3HK*,QA#,KA# |
| | | | | | 23<br>0/23 | | 52<br>0/52 | 31.8(6.3) | | | | | | | TRP*,KYN*,<br>3HK$^{\#}$,QA*,KA* |
| | | | | | 29<br>0/29 | | 58<br>0/58 | 32.2(5.4) | | | | | | | TRP*,KYN$^{\#}$,<br>3HK$^{\#}$,QA*,KA* |
| | | | | | 29<br>0/29 | | 58<br>0/58 | 33.02(4.6) | | | | | | | TRP*,KYN$^{\#}$,<br>3HK$^{\#}$,QA*,KA* |
| 112 | Wood, Harwood et al. 1978 | UK | MDD | Healthy Control | 10<br>10/0 | 14<br>14/0 | 24<br>24/0 | 55,9(15,2) | 53,1(10,8) | KYN | Plasma | Flame photomet<br>ry | 4 | 15,5 | KYN$^{\#}$ |
| | | | | | 17 | 16 | 33 | 59,6(9,7) | 52,5(8,5) | | | | | | KYN* |

| | | | | | | | | | | | | | | | |
|---|---|---|---|---|---|---|---|---|---|---|---|---|---|---|---|
| | | | | | 0/17 | 0/16 | 0/33 | | | | | | | | |
| 113 | Wu, Mai et al. 2018a | China | MDD | Healthy Control | 170<br>43/127 | 135<br>42/93 | 305<br>85/220 | 66,5(7,01) | 66,9(6,7) | TRP, KYN, KA | Serum | LC-MS/MS | 6,75 | 12,5 | TRP*, KYN*, KA* |
| 114 | Wu, Zhong et al. 2018b | China | MDD | Healthy Control | 85<br>20/65 | 129<br>38/91 | 214<br>58/156 | 66,3(7,3) | 67,08(6,8) | TRP, KYN, KA | Serum | LC-MS/MS | 5,75 | 14 | TRP*, KYN*, KA* |
| | | | | | 71<br>22/49 | | 200<br>71/129 | 66,7(6,9) | | | | | | | TRP*, KYN*, KA* |
| 115 | Wurfel, Drevets et al. 2017 | USA | MDD | Healthy Control | 35<br>16/19 | 92<br>33/59 | 127<br>49/78 | 38.8(13.8) | 32.3(10.4) | TRP,KYN, KA,3HK,QA | Serum | HPLC-MS/MS | 3,75 | 20 | TRP*,KYN, KA*,3HK*,QA[#] |
| | | | BD | | 53<br>16/37 | | 145<br>49/96 | 40.2(11.0) | | | | | | | TRP*,KYN*, KA*,3HK*,QA[#] |
| 116 | Xu, Fang et al. 2012 | China | MDD | Healthy Control | 26<br>7/19 | 25<br>9/16 | 51<br>16/35 | 32.3(8.8) | 32.1(8.1) | TRP | Plasma | HPLC | 7 | 9,5 | TRP* |
| 117 | Chiu, Yang et al. 2021 | Taiwan | MDD | Healthy Control | 33<br>8/25 | 33<br>8/25 | 66<br>16/50 | 43.7(11.6) | 43.3(11.3) | TRP, KYN | Plasma | ELISA | 5,75 | 12,5 | TRP*, KYN[#] |
| 118 | Yoshimi, Futamura et al. 2016 | Sweden | BD | Healthy Control | 54<br>54/0 | 39<br>39/0 | 93<br>93/0 | 41.7(15.2) | 37.4(13.8) | TRP | Serum | CE-TOFMS | 5 | 20 | TRP* |
| 119 | Young, Drevets et al. 2016 | USA | MDD | Healthy Control | 54<br>54/0 | 39<br>39/0 | 93<br>93/0 | 41.7(15.2) | 38.1(15.3) | TRP,KYN, KA,3HK, QA | Serum | CE-TOFMS | 8,75 | 9,5 | TRP*,KYN, KA*,3HK[#], QA[#] |
| 120 | Zhou, Zheng et al. 2018 | China | MDD | Healthy Control | 84<br>39/45 | 60<br>35/25 | 144<br>74/70 | 35.0(12.1) | 31.6(10.7) | TRP,KYN, KA | Serum | HPLC-MS/MS | 5 | 12,5 | TRP*,KYN*, KA* |
| 121 | Zhou, Zheng et al. 2019 | China | MDD | Healthy Control | 76<br>76/0 | 41<br>41/0 | 117<br>117/0 | 33,3(11,7) | 28,8(9,7) | TRP,KYN, KA | Serum | HPLC-MS/MS | 4,5 | 17,5 | TRP*,KYN*, KA* |
| | | | | | 70<br>0/70 | 31<br>0/31 | 101<br>0/101 | 35(12,4) | 36,3(11,9) | | | | | | TRP*,KYN[#], KA* |
| 29<br>USA | UK<br>10 | China<br>8 | 8<br>Germany | 7<br>Belgium | 6<br>Japan | 6<br>Netherla nd | 6<br>Italy | 5<br>Austria | 4<br>Ireland | 3<br>Denemark | 9<br>Sweden | | | | |

*: Indicates that patients have reduced level of the measured metabolite compared to healthy control

#: Indicates that patients have increased level of the measured metabolites compared to healthy control

TRP: Tryptophan, KYN: Kynurenine, KA: Kynurenic acid, 3HK: 3-Hydroxykynurenine, AA: Anthranilic acid, 3HA: 3-Hydroxyanthranilic acid, XA: Xanthurenic acid, QA: Quinolinic acid, PA: Picolinic acid, MDD: Major depressive disorder, BD: Bipolar disorder, MDD-M: Major depressive disorder with melancholia, DD: Dysthymic disorder, UD: Unipolar depression, HPLC: High performance liquid chromatography, HPLC-MS/MS: High performance liquid chromatography with tandem mass spectrometry, CE-TOFMS: Capillary Electrophoresis Time Of Flight Mass Spectrometer, LC-MS/MS: Liquid chromatography with tandem mass spectrometry, UPLC-MS/MS: Ultra performance liquid chromatography with tandem mass spectrometry, LCECA: Liquid, chromatography electrochemical array, XLC-MS/MS: Extraction-liquid chromatographic-tandem mass spectrometric, GCMS: Gas chromatography–mass spectrometry, LC: Liquid chromatography, HPLC-UV: High perfomance liquid chromatography- Ultra-violate

**ESF. Table 6**. Results of Meta-regression

| Variables | No. of Studies | Covariates | 1-sided p-value | Z-Value |
|---|---|---|---|---|
| TRP | 80 | Medication Free | 0.006 | -2.48 |
| | 104 | Ogyu2018 | 0.011 | 2.26 |
| | | Ogawa2014 | 0.044 | -1.70 |
| | 83 | Sample size | 0.032 | 1.84 |
| | 79 | Male cases | 0.047 | 1.67 |
| TRP/CAAs | 13 | Total Cases | 0.008 | 2.38 |
| CAAs | 14 | Latitude | 0.016 | 2.15 |
| | 10 | Partially Free Medication | 0.021 | 2.03 |
| | | Medication Free | 0.019 | 2.06 |
| (KYN+3HK+3HA+XA+QA+PA) | 82 | Bartoli-2021 | 0.002 | -2.87 |
| | | Marx-2020 | 0.010 | -2.31 |
| | | Plasma | 0.001 | -3.03 |
| | | Serum | 0.027 | -1.93 |
| KA/KYN | 69 | ECT-Yes | 0.043 | -1.71 |

| | | | | |
|---|---|---|---|---|
| | 67 | Ogyu-2018 | 0.025 | -1.96 |
| | | Marx-2020 | 0.033 | -1.84 |
| | 41 | Severity-Moderate | 0.039 | 1.76 |
| KA | 32 | Severity-Moderate | 0.0012 | 3.04 |
| | 55 | Ogyu-2018 | 0.013 | -2.22 |
| | | Marx-2020 | 0.0004 | -3.34 |
| | | Hebbrecht-2021 | 0.0061 | -2.50 |
| 3HK | 21 | Age | 0.012 | 2.24 |

TRP: Tryptophan, KYN: Kynurenine, KA: Kynurenic acid, 3HK: 3-Hydroxykynurenine, AA: Anthranilic acid, 3HA: 3-Hydroxyanthranilic acid, XA: Xanthurenic acid, QA: Quinolinic acid, PA: Picolinic acid, MDD: CAAs: Competing amino acids.

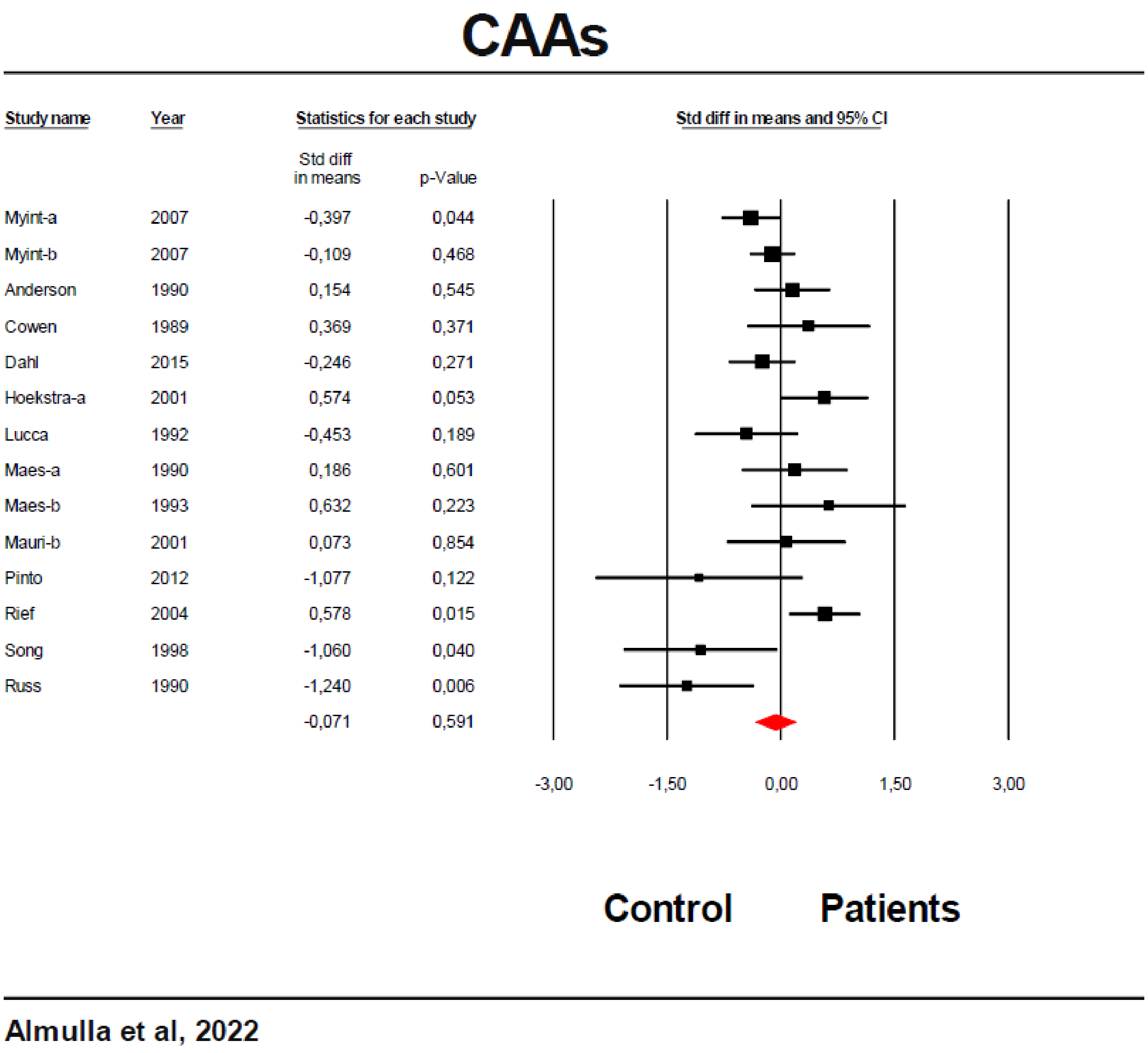

**ESF, Figure 1**. Forest plot of competing amino acids (CAAs) in the patients with affective disorders versus healthy control.

### TRP/CAAs

| Study name | | Std diff in means | p-Value |
|---|---|---|---|
| DeMyer | 1981 | -1,508 | 0,001 |
| Cowen | 1989 | -0,576 | 0,189 |
| Anderson | 1990 | -0,280 | 0,278 |
| Maes-a | 1990 | -0,263 | 0,467 |
| Russ | 1990 | -1,190 | 0,008 |
| Lucca | 1992 | -0,958 | 0,008 |
| Maes-b | 1993 | -0,484 | 0,379 |
| Moller-a | 1993 | -0,662 | 0,007 |
| Maes-e | 1996 | -1,042 | 0,006 |
| Maes-g | 1997 | -1,017 | 0,002 |
| Song | 1998 | -0,900 | 0,080 |
| Hoekstra-a | 2001 | -0,394 | 0,202 |
| Mauri-b | 2001 | -0,041 | 0,918 |
| Rief | 2004 | -0,229 | 0,346 |
| Myint-a | 2007 | -0,543 | 0,006 |
| Myint-b | 2007 | -0,193 | 0,200 |
| Pinto | 2012 | -0,914 | 0,182 |
| Dahl | 2015 | -0,038 | 0,864 |
| | | -0,522 | 0,000 |

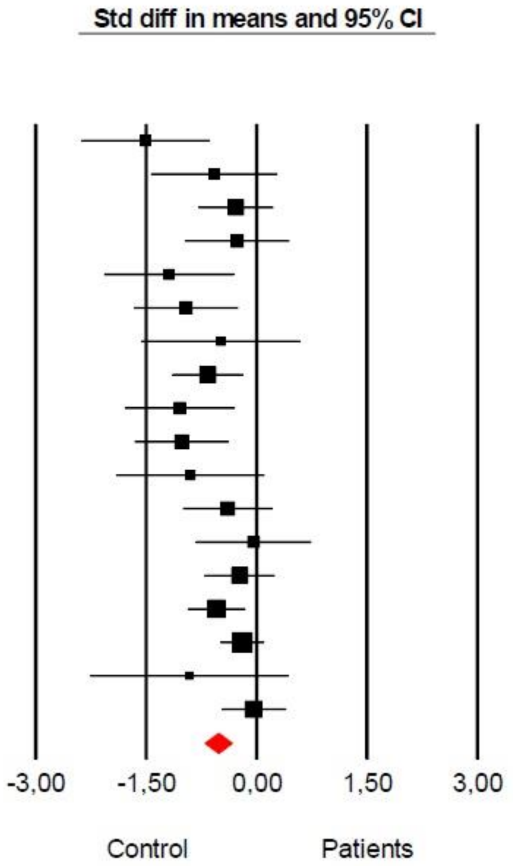

**Almulla et al, 2022**

**ESF, Figure 2**. Forest plot of tryptophan (TRP)/competing amino acids (CAAs) in the patients with affective disorders versus healthy control.

**Abbreviation:**

TRP: Tryptophan

KYN: Kynurenine

KA: Kynurenic acid

3HK: 3-Hydroxykynurenine

AA: Anthranilic acid

3HA: 3-Hydroxyanthranilic acid

XA: Xanthurenic acid

QA: Quinolinic acid

PA: Picolinic acid

CAAs: Competing amino acids

IDO: Indoleamine 2,3 dioxygenase

TDO: Tryptophan 2,3 dioxygenase

KAT: Kynurenine aminotransferase

KMO: Kynurenine 3-monooxygenase

KYNU: Kynureninase

$NAD^+$: Nicotinamide adenine dinucleotide

TRYCATs: Tryptophan Catabolites

TRYCAT pathway: Tryptophan catabolite pathway

SMD: Standardized mean difference

IL-6: Interleukin-6

IFN-γ: Interferon-gamma

O&NS: Oxidative and nitrosative stress

NAD: Nicotinamide adenine dinucleotide

MOOSE: Meta-Analyses of Observational Studies in Epidemiology

CSF: Cerebrospinal fluid

SD: Standard definition

IOR: Interquartile range

ICS: Immune confounder scale

CI: Confidence intervals

LC-MS: Liquid chromatography-mass spectrometry

LC-MS/MS: Liquid chromatography with two mass spectrometry

UHPLC-MS: Ultra-high-performance liquid-chromatography- mass spectrometry

LC-HRMS: Liquid chromatography–high-resolution mass spectrometry

LC-UV: Liquid chromatography-UV detection

NMDA: N-methyl-D-aspartate
